# Trends in daily cigarette consumption among smokers: a population study in England, 2008-2023

**DOI:** 10.1101/2023.12.05.23299431

**Authors:** Sarah E. Jackson, Harry Tattan-Birch, Vera Buss, Lion Shahab, Jamie Brown

**Affiliations:** Department of Behavioural Science and Health, University College London, London, UK; SPECTRUM Consortium, UK

**Keywords:** cigarette consumption, cigarettes per day, time trends, population study, roll-your-own

## Abstract

**Objectives:** To estimate time trends in cigarette consumption among smokers in England between 2008 and 2023, and explore differences by key potential moderators.

**Methods:** We used data from 57,778 adult cigarette smokers participating in a nationally-representative monthly cross-sectional survey between January-2008 and September-2023. We estimated monthly time trends in mean daily consumption of (i) any, (ii) manufactured, and (iii) hand-rolled cigarettes among all smokers and by main type of cigarettes smoked, smoking frequency, age, gender, occupational social grade, region, nicotine replacement therapy use, and vaping status.

**Results:** Overall cigarette consumption fell from 13.6 [95%CI=13.3-13.9] to 10.6 [10.5-10.8] per day between January-2008 and October-2019 (a 22% decrease), then remained stable up to September-2023. Over this period, the proportion mainly/exclusively smoking hand-rolled cigarettes increased (from 30.6% [29.1-32.1%] in 2008 to 52.1% [49.7-54.5%] in 2023). As a result, manufactured cigarette consumption fell by 47%, from 9.5 [9.2-9.8] per day in January-2008 to 5.0 [4.7-5.3] in September-2023, while hand-rolled cigarette consumption increased by 35%, from 4.2 [3.9-4.4] to 5.6 [5.3-5.9]. The decline in overall cigarette consumption was observed across all subgroups, but was greater among non-daily smokers, younger smokers, and those who vaped.

**Conclusions:** Over the last 15 years, the average number of cigarettes consumed each day by smokers in England has fallen by almost a quarter, but has plateaued since October-2019. There has been a sharp decline in the number of manufactured cigarettes consumed and an increase in the number of hand-rolled cigarettes consumed, as smokers have increasingly shifted towards using hand-rolled tobacco.

## Introduction

Cigarettes are uniquely lethal, eventually killing up to two thirds of people who smoke throughout their lives.^1,2^ While there is no safe level of consumption,^3,4^ the risks are greater for people who smoke more cigarettes per day.^3–10^ In England, the proportion of adults who smoke cigarettes has declined steadily over recent decades.^11^ However, a substantial minority (12.7% in 2022^11^) continue to smoke, with higher rates among certain population groups.^11^ It is important to understand the extent to which smokers’ daily cigarette consumption is changing over time at a national level, and how these changes vary between groups. This information can inform estimates of smoking harms and evaluation of policies aiming to reduce smoking and associated inequalities in health.

Daily cigarette consumption is an important determinant of health outcomes. Considerable evidence has shown associations between the number of cigarettes smoked per day and the risk of developing or dying from smoking-related diseases.^3–10^ These risks are not necessarily linear,^3^ although some of the non-linear effects that have been documented may reflect the fact that self-reported daily cigarette consumption is not a perfect marker for exposure, given that individuals may get more or less nicotine (and with it, harmful substances) out of each cigarette they smoke.^12,13^ On a population level, there is evidence that reducing per-capita tobacco consumption is associated with lower rates of death from heart disease and cancer.^14–17^ On an individual level, there is also some evidence that reducing the amount smoked can lower these risks.^18^ In addition to its direct associations with morbidity and mortality, the risks of higher cigarette consumption may be compounded by a possible association with lower odds of future quitting. Several studies suggest people who smoke more heavily are less inclined to try to quit or have less success in quitting compared with people who smoke fewer cigarettes.^19–22^ There is mixed evidence as to whether reducing the number of cigarettes smoked per day increases smokers’ odds of making a quit attempt or successfully quitting, with evidence that reduction is more effective when supported by varenicline or nicotine replacement therapy (NRT).^23–28^

Data from representative population surveys in England have shown a decline in cigarette consumption over recent years. The Health Survey for England (HSE) recorded a fall in the median number of cigarettes smoked per day among adult smokers aged ≥16 years, from 15 cigarettes per day in 1993 to 9 cigarettes per day in 2021.^29^ Similarly, in the Opinions and Lifestyle Survey (OPN), mean daily cigarette consumption among adult smokers aged ≥16 years fell from 14 in 2000 to 11 in 2017 (and to 9 in 2019, although the question was worded differently, meaning this estimate is not directly comparable to previous years).^30^ The Smoking Toolkit Study estimated that population-level adult cigarette consumption fell by 118.4 million cigarettes per month between 2011 and 2018.^31^ Cigarette sales data over this period corroborated this decline.^31^ However, as an estimate of change in total cigarette consumption in the population, this number reflects a reduction in the total number of smokers as well as in the number of cigarettes consumed by those who smoke.

Little is known about how these changes in cigarette consumption have varied across subgroups of smokers. HSE and OPN report data on cigarettes per day separately by sex, which suggest the decline has been similar among men and women.^29,30^ However, it is possible that other factors may be more strongly linked to trends in cigarette consumption. One important influence on cigarette consumption is affordability.^32,33^ Therefore, changes in consumption may vary according to the type of cigarettes people smoke (with hand-rolled cigarettes substantially cheaper than manufactured cigarettes^34^) or their disposable income (which tends to be lower among younger people^35^ and those working in routine and manual occupations). There may also be age or regional differences in changes in cigarette consumption.^36,37^ In addition, since e-cigarettes have become popular, changes in cigarette consumption may also have differed between smokers who do and do not also vape, given many dual users (of cigarettes and e-cigarettes) report vaping to reduce the amount they smoke.^38^

This study aimed to estimate time trends in average daily cigarette consumption among adult current smokers in England between 2008 and 2023, and explore differences by key potential moderators. Specific research questions were:

1. How has current smokers’ mean daily consumption of (a) any cigarettes, (b) manufactured cigarettes, and (c) hand-rolled cigarettes changed since 2008?
2. To what extent have these changes differed by the main type of cigarettes smoked (manufactured/hand-rolled), frequency of smoking (daily/non-daily), age, gender, occupational social grade, region in England, use of NRT, and vaping status?

## Methods

### Pre-registration

The study protocol and analysis plan were pre-registered on Open Science Framework (https://osf.io/bg7dv/) and followed without amendment. In addition to our planned analyses, we presented descriptive data on the annual proportions of current smokers (i) using manufactured vs. hand-rolled cigarettes and (ii) smoking daily vs. non-daily, to provide additional context.

### Design

Data were drawn from the ongoing Smoking Toolkit Study, a monthly cross-sectional survey of a representative sample of adults aged ≥16 years in England.^39^ The study uses a hybrid of random probability and simple quota sampling to select a new sample of approximately 1,700 adults each month. Comparisons with sales data and other national surveys indicate that key variables including sociodemographic characteristics, smoking prevalence, and cigarette consumption are nationally representative.^31,39^

We used data collected from January 2008 (the first year to collect data on the type of cigarettes smoked) through September 2023 (the most recent data at the time of analysis). Data were initially collected through face-to-face computer-assisted interviews. However, social distancing restrictions under the Covid-19 pandemic meant no data were collected in March 2020 and data from April 2020 onwards were collected via telephone. The telephone-based data collection used the same combination of random location and quota sampling, and weighting approach as the face-to-face interviews and comparisons of the two data collection modalities indicate good comparability.^40–42^

For analysis, we selected respondents who reported current cigarette smoking. Because data were not collected from 16- and 17-year-olds between April 2020 and December 2021, we restricted our sample to those aged ≥18 for consistency across the time series.

### Measures

***Smoking status*** was assessed by asking participants which of the following best applied to them:

a. ‘I smoke cigarettes (including hand-rolled) every day’
b. ‘I smoke cigarettes (including hand-rolled), but not every day’
c. ‘I do not smoke cigarettes at all, but I do smoke tobacco of some kind (e.g. pipe, cigar or shisha)’
d. ‘I have stopped smoking completely in the last year’
e. ‘I stopped smoking completely more than a year ago’
f. ‘I have never been a smoker (i.e. smoked for a year or more)’

Those who responded *a* or *b* were considered current cigarette smokers. Those who responded *c* to *f* were excluded from the analytic sample.

***Cigarette consumption*** was assessed with the questions: ‘How many cigarettes do you usually smoke?’ and ‘How many of these do you think are hand-rolled?’. Participants who responded ‘don’t know’ were encouraged to give their best estimates. Participants could choose to give their answers per day or per week (or per month, in some waves). Our primary outcome was total daily cigarette consumption (i.e., cigarettes per day; or cigarettes per week divided by 7 or per month divided by 30). Secondary outcomes were daily hand-rolled cigarette consumption and daily manufactured cigarette consumption (calculated as total daily cigarette consumption minus daily hand-rolled cigarette consumption). These were analysed as continuous variables. We considered consumption of more than 80 cigarettes per day to be implausible,^43^ so replaced responses above this level with the value 80.

***Main type of cigarettes smoked*** was defined as hand-rolled for those who reported at least 50% of their total cigarette consumption is hand-rolled, and manufactured for those who reported that less than 50% is hand-rolled. This definition has been used in previous studies^44–46^ and allows inclusion of those who smoke both hand-rolled and manufactured cigarettes as well as those who exclusively smoke one type of cigarettes.

***Frequency of smoking*** was defined according to responses to the question assessing smoking status (described above), with those who responded *a* considered daily cigarette smokers and those who responded *b* non-daily cigarette smokers.

***Age*** was analysed as a continuous variable, modelled using restricted cubic splines (see *Statistical analysis* section). We also provide descriptive data by age group (18-24/25-34/35-44/45-54/55-64/≥65).

***Gender*** was self-reported as man or woman. In more recent waves, participants have also had the option to describe their gender in another way; those who identified in another way were excluded from analyses by gender due to low numbers.

***Occupational social grade*** was categorised as ABC1 (includes managerial, professional, and upper supervisory occupations) and C2DE (includes manual routine, semi-routine, lower supervisory, and long-term unemployed).

***Region in England*** was categorised as north, midlands, and south.

***NRT use*** was assessed with a series of questions that ask participants whether they are using a range of nicotine products to help them stop smoking, cut down the amount smoked, in situations when smoking is not permitted, or for any other reason at all. Those who reported using NRT (nicotine gum, lozenges/tablets, inhaler, nasal spray, patch, or mouth spray) in response to any of these questions were considered current NRT users.

***Vaping status*** was assessed with the same series of questions used to assess NRT use. Those who reported using an e-cigarette in response to any of these questions were considered current vapers. Data on current e-cigarette use were only available from April 2011, so analyses by vaping status were limited to this period. Frequency of vaping was assessed with the question: ‘How many times per day on average do you use your nicotine replacement product or products?’ Our primary analysis defined vaping status as any current use of e-cigarettes (yes/no); a sensitivity analysis compared daily and non-daily vapers.

### Statistical analysis

Analyses were done using R v.4.2.1. The Smoking Toolkit Study uses raking to weight the sample to match the population in England. This profile is determined each month by combining data from the UK Census, the Office for National Statistics mid-year estimates, and the annual National Readership Survey.^39^ The following analyses used weighted data. We excluded participants with missing data on overall daily cigarette consumption. Missing cases on other variables (see **Table S1** for details) were excluded on a per-analysis basis.

We reported descriptive data on daily (i) overall, (ii) manufactured and (iii) hand-rolled cigarette consumption by survey year, among all adult cigarette smokers and within subgroups.

We used linear regression to estimate monthly time trends in daily (i) overall, (ii) manufactured and (iii) hand-rolled cigarette consumption across the study period. Time (survey wave) was modelled using restricted cubic splines with five knots (sufficient to accurately model trends across years without overfitting), to allow relationships with time to be flexible and non-linear, while avoiding categorisation.

To explore moderation by main type of cigarettes smoked, frequency of smoking, age, gender, occupational social grade, region in England, NRT use, and vaping status, we repeated the models including the interaction between the moderator of interest and time – thus allowing for time trends to differ across sub-groups. Each of the interactions was tested in a separate model. Age was modelled using restricted cubic splines with three knots (placed at the 5, 50, and 95% quantiles), to allow for a non-linear relationship between age and daily cigarette consumption. We conducted a sensitivity analysis within current vapers comparing trends among daily vs. non-daily users (i.e., testing the interaction between frequency of vaping [daily/non-daily] and time).

We used predicted estimates from these models to (i) plot mean daily cigarette consumption over the study period and (ii) derive specific predictions for current levels of cigarette consumption (in September 2023), among all cigarette smokers and within each subgroup of interest, incorporating information from all survey months and thus increasing statistical precision. As age was modelled continuously, we displayed estimates for six specific ages (18-, 25-, 35-, 45-, 55-, and 65-year-olds) to illustrate how trends differ across ages. Note that the model used to derive these estimates included data from participants of all ages, not only those aged exactly 18, 25, 35, 45, 55, or 65 years.

## Results

A total of 318,083 (unweighted) adults aged ≥18 years were surveyed between January 2008 and September 2023, of whom 58,974 (18.5%) reported current cigarette smoking. We excluded 1,196 (2.0%) with missing data on daily cigarette consumption, leaving a final sample for analysis of 57,778 participants. Forty-four participants (0.08%) reported smoking more than 80 cigarettes per day; their consumption was imputed as 80 in line with our assumptions around plausibility (see *Measures* section). Sample characteristics are summarised in **Table S1**.

### Trends in cigarette consumption among all current smokers

Among all current smokers, mean overall cigarette consumption decreased from 13.5 [95%CI 13.2-13.7] cigarettes per day in 2008 to 11.0 [10.5-11.4] in 2023 (**Table S2; Figure 1A**). Our model indicated this decline was non-linear (**Figure 1B**), falling steadily between January 2008 and October 2019, from 13.6 [13.3-13.9] to 10.6 [10.5-10.8], then remaining stable between 10.5 and 10.6 up to September 2023. As the mean number of cigarettes smoked per day declined, the distribution became more skewed, with an increasing proportion of smokers reporting low levels of consumption (<5 cigarettes per day; **Figure 2**) and smoking non-daily (**Figure S1**). In addition, there was a shift in the main type of cigarettes people were smoking: the proportion mainly smoking hand-rolled cigarettes increased from 30.6% [29.1-32.1%] in 2008 to 52.1% [49.7-54.5%] in 2023 (**Figure S2**). As a result, mean consumption of manufactured cigarettes fell from 9.3 [9.0-9.6] per day in 2008 to 5.4 [5.0-5.8] in 2023 (**Table S3**), while mean consumption of hand-rolled cigarettes increased from 4.2 [4.0-4.4] to 5.7 [5.3-6.1] per day (**Table S4; Figure 1A**). These changes occurred predominantly between 2008 and 2015, with the rate of decline in manufactured cigarette consumption and the rate of increase in hand-rolled cigarette consumption slowing notably from 2016 onwards (**Figure 1B**).

**Figure 1.**
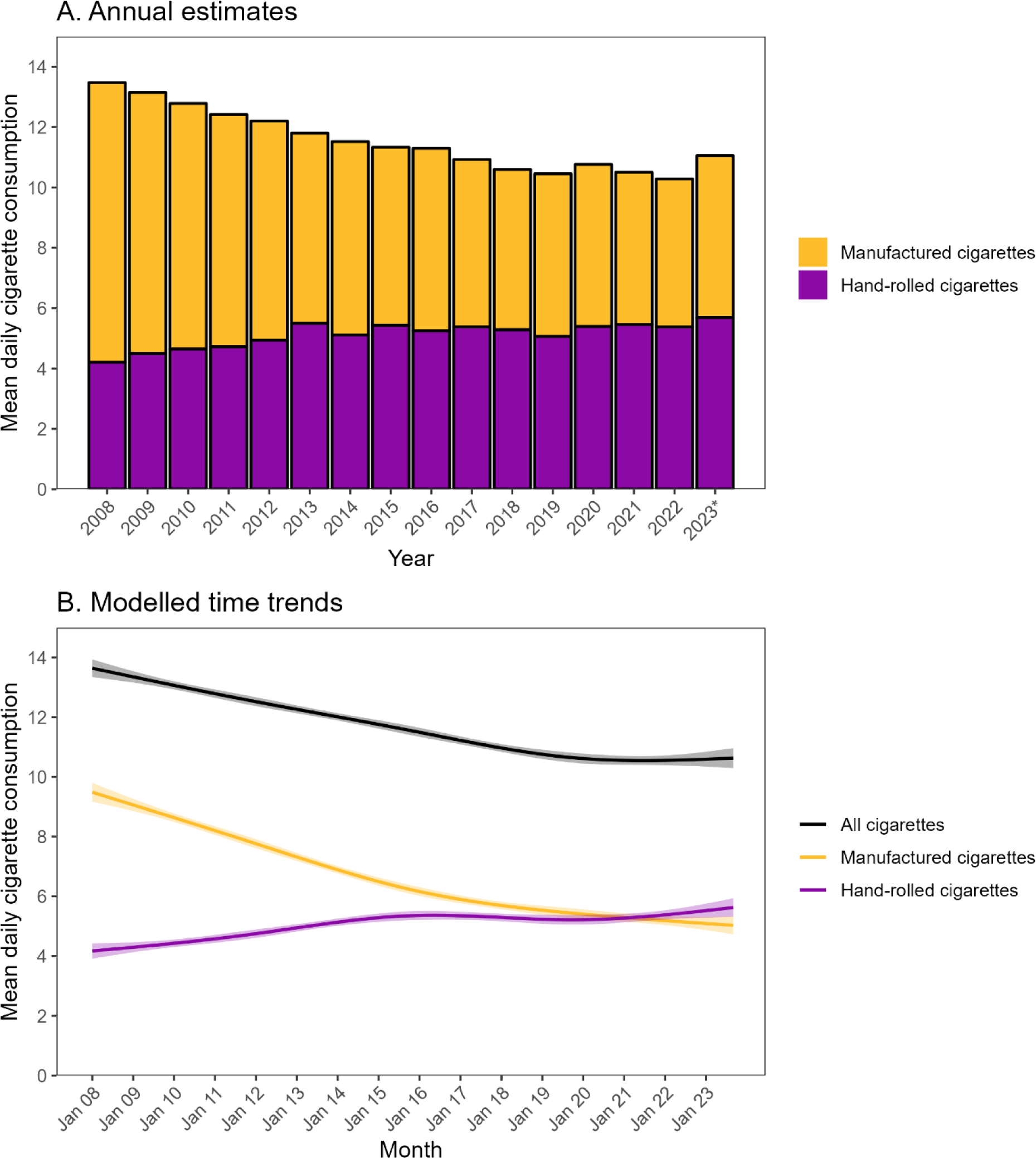
Mean daily cigarette consumption among adult (≥18y) smokers in England, 2008 to 2023. Panel A shows weighted data aggregated by year. Bars represent mean overall daily cigarette consumption as a function of cigarette type (manufactured vs. hand-rolled). * Data for 2023 are based on January to September only. *Annual estimates of mean daily cigarette consumption within each subgroup of interest are provided in Tables S2 (all cigarettes), S3 (manufactured cigarettes), and S4 (hand-rolled cigarettes)*. Panel B shows modelled time trends, overall and by cigarette type. Lines represent modelled weighted mean daily cigarette consumption by monthly survey wave, modelled non-linearly using restricted cubic splines (five knots). Shaded bands represent 95% confidence intervals. *Modelled estimates of mean daily cigarette consumption (overall and by cigarette type) at the start and end of the study period, among all smokers and within each subgroup of interest, are provided in Table 1*.

**Figure 2.**
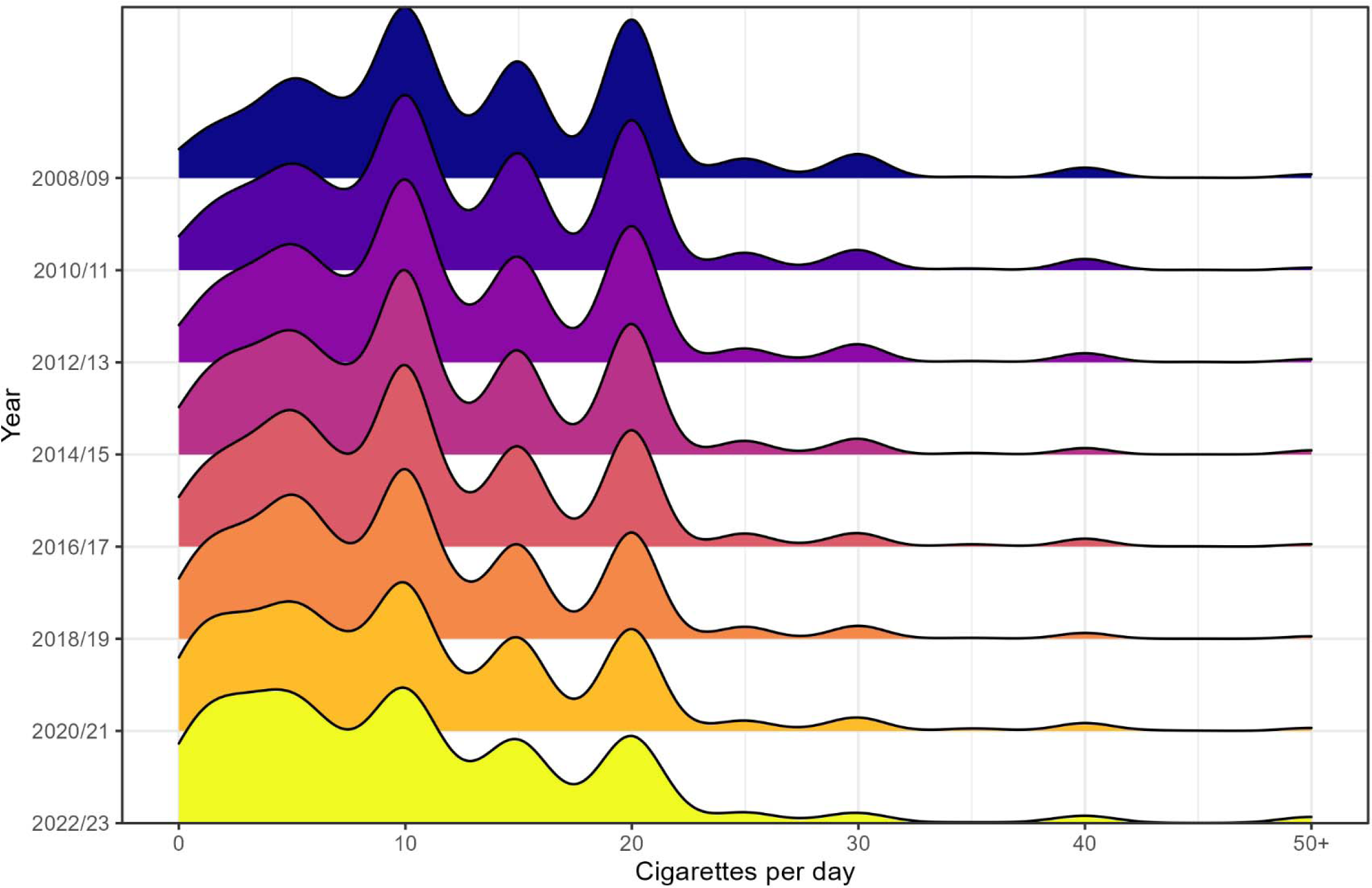
Distributions of overall daily cigarette consumption among adult (≥18y) smokers in England, 2008/09 to 2022/23.

**Table 1.**
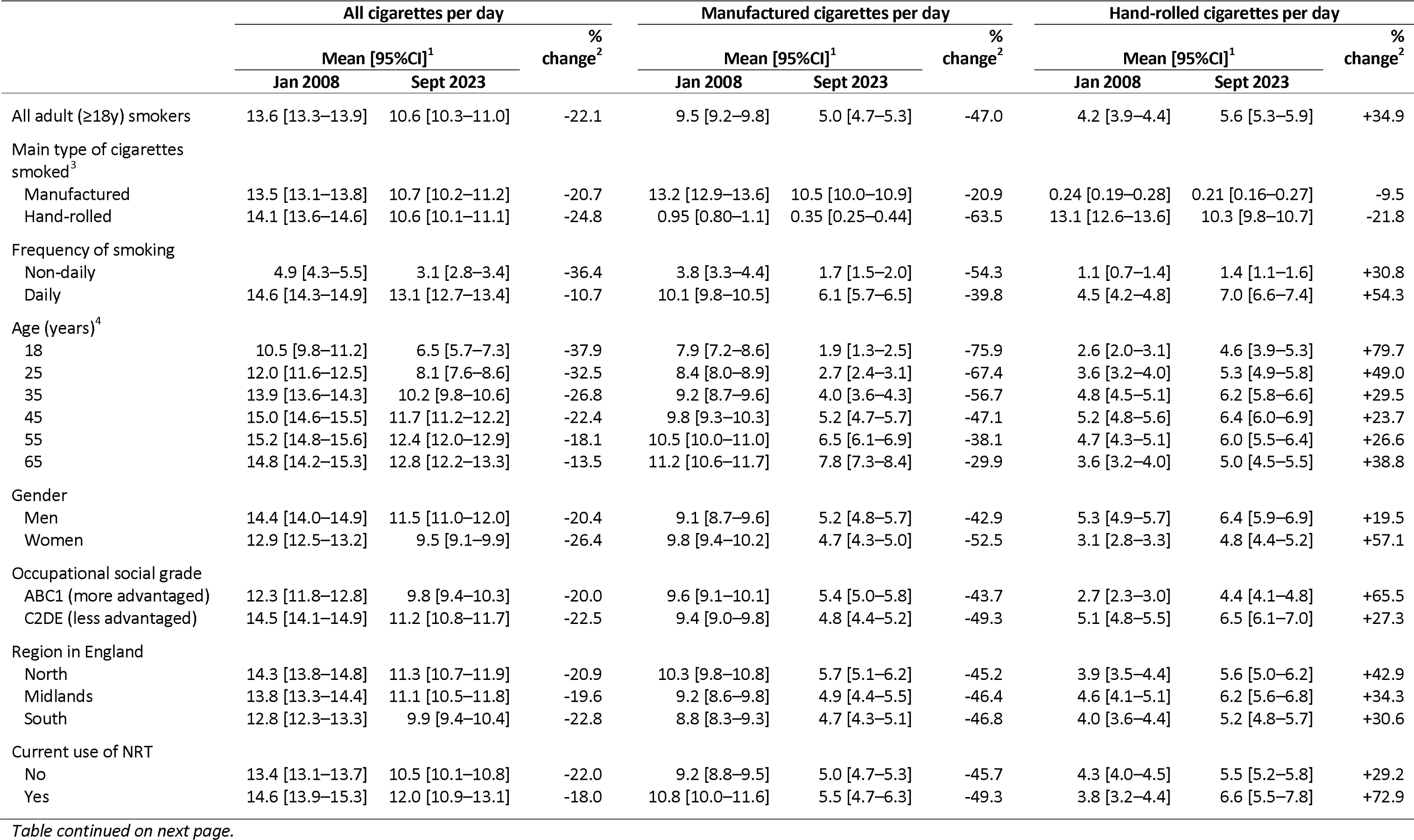
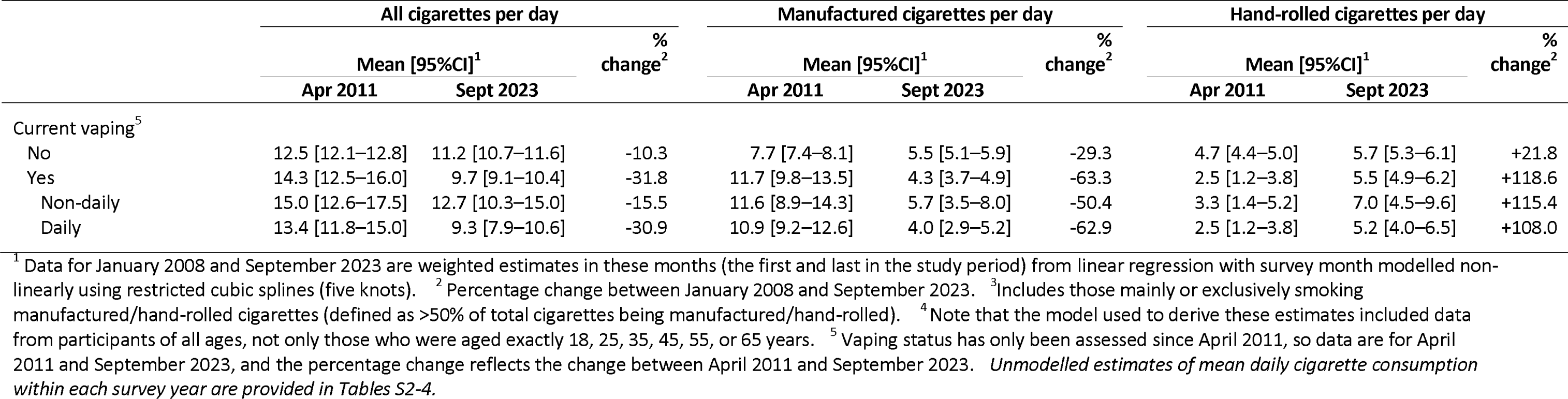
Modelled estimates of the change in daily cigarette consumption among adult smokers (≥18y) in England from January 2008 to September 2023.

### Trends in cigarette consumption within subgroups of smokers

Across the study period, mean overall cigarette consumption was consistently higher among daily smokers and those who were older, men, from more disadvantaged social grades (C2DE), and living in the north (vs. south) of England (**Table S2; Figure 3**). Mean manufactured cigarette consumption was consistently higher among those who mainly or exclusively used manufactured cigarettes, daily smokers, and those who were older and living in the north (vs. south) of England (**Table S3; Figure S3**) and mean hand-rolled cigarette consumption was consistently higher among those who mainly or exclusively used hand-rolled cigarettes, daily smokers, men, and those from more disadvantaged social grades (C2DE; **Table S4; Figure S4**).

**Figure 3.**
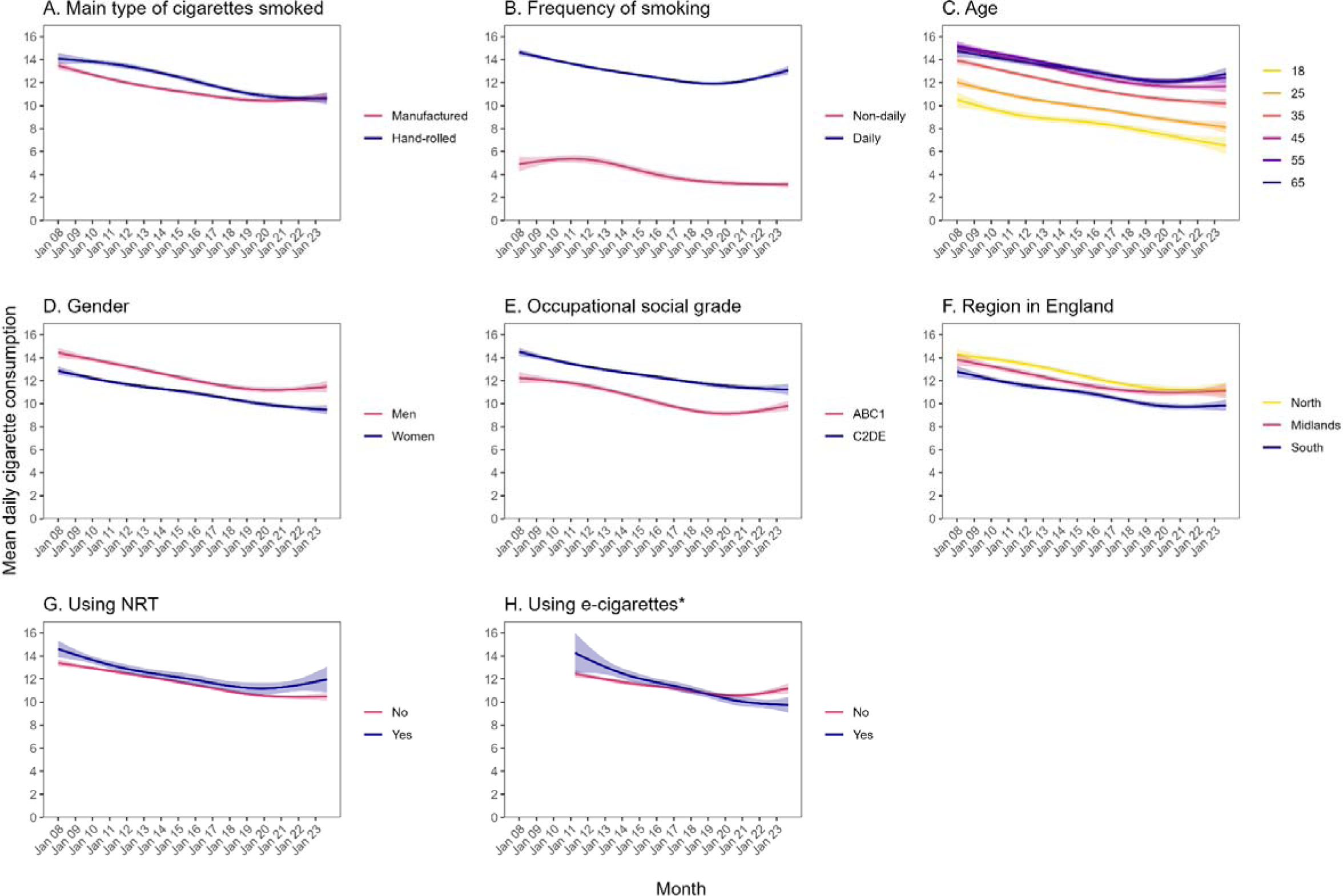
Time trends in overall daily cigarette consumption among adult (≥18y) smokers in England, January 2008 to September 2023. Panels show trends by (A) main type of cigarettes smoked (includes those mainly or exclusively smoking manufactured/hand-rolled cigarettes, defined as >50% of total cigarettes), (B) frequency of smoking, (C) age, (D) gender, (E) occupational social grade, (F) region in England, (G) NRT use, and (H) vaping status. Lines represent modelled weighted mean daily cigarette consumption by monthly survey wave, modelled non-linearly using restricted cubic splines (five knots). Shaded bands represent 95% confidence intervals. * Vaping status has only been assessed since April 2011. *Corresponding figures for daily consumption of manufactured and hand-rolled cigarettes are provided in Figures S3 and S4*.

From the start to the end of the study period, we observed a significant decline in overall cigarette consumption across all subgroups (**Table 1**; **Figure 3**). The relative extent of this decline was greater among non-daily smokers (-36.4% vs. -10.7% among daily smokers), those who were younger (e.g., -37.9% among 18-year-olds vs. -13.5% among 65-year-olds), and those who currently vaped (-31.8% vs. -10.3% among non-vapers) – particularly those who vaped daily (-30.9% vs. -15.5% among non-daily vapers; **Figure S5**). The association between vaping status and overall cigarette consumption reversed during the course of the study, with smokers who vaped reporting slightly higher overall cigarette consumption in April 2011 (the first wave in which vaping status was assessed; 14.3 vs. 12.5 cigarettes per day), but significantly lower consumption by the end of the study (September 2023; 9.7 vs. 11.2; **Table 1**, **Figure 3**). The decline in overall cigarette consumption appeared to level off, or possibly reverse, from 2019 onwards more prominently among those who were daily smokers, aged ≥55, men, from more advantaged social grades (ABC1), and did not currently vape (**Figure 3**). The plateau was more consistent within the remaining subgroups.

Manufactured cigarette consumption also declined significantly across all subgroups (**Table 1; Figure S3**). The relative extent of this decline was greater among those who mainly or exclusively smoked hand-rolled cigarettes (-63.5% vs. -20.9% among those who mainly or exclusively smoked manufactured cigarettes), non-daily smokers (-54.3% vs. -39.8% among daily smokers), those who were younger (e.g., -75.9% among 18-year-olds vs. -29.9% among 65-year-olds), women (-52.5% vs. -42.9% among men), and those who currently vaped (-63.3% vs. -29.3% among non-users). As we observed for overall consumption, manufactured cigarette consumption was significantly higher among smokers who vaped at the start of the study but significantly lower by the end.

Hand-rolled cigarette consumption increased significantly across most, but not all, subgroups (**Table 1; Figure S4**). Notably, the number of hand-rolled cigarettes smoked per day *declined* by 21.8% among those who mainly or exclusively smoked hand-rolled cigarettes and did not change significantly among those who mainly or exclusively smoked manufactured cigarettes. This suggests the increase we observed among all smokers and within other subgroups was the result of people switching from manufactured to hand-rolled cigarettes, rather than people who smoked hand-rolled cigarettes increasing their consumption. Consistent with this, we saw an increase in the *proportion* who mainly or exclusively smoked hand-rolled cigarettes across the study period within each of the other subgroups of interest (**Figure S6**). In addition, most of the subgroups with a greater relative decline in manufactured cigarette consumption also showed a greater relative increase in hand-rolled consumption: those who were younger (e.g., +79.7% among 18-year-olds vs. +38.8% among 65-year-olds), women (+57.1% vs. +19.5% among men), and those who currently vaped (+118.6% vs. +21.8% among non-users). The increase in hand-rolled cigarette consumption was also greater among daily smokers (+54.3% vs. +30.8% among non-daily smokers), those from more advantaged social grades (ABC1; +65.5% vs. 27.3% among C2DE), and those using NRT (+72.9 vs. +29.2% among non-users).

## Discussion

Between January 2008 and September 2023, the average number of cigarettes consumed by adult smokers in England decreased non-linearly from 13.6 to 10.6 per day. At the same time, the proportion mainly or exclusively smoking hand-rolled cigarettes increased substantially. This saw the average number of manufactured cigarettes smokers consumed per day fall from 9.5 to 5.0, while the number of hand-rolled cigarettes consumed increased from 4.2 to 5.6. The decline in overall cigarette consumption was observed across all subgroups, but was greater among non-daily smokers, those who were younger, and those who currently vaped.

Annual surveys have documented a decline in the average number of cigarettes consumed per day by smokers in England.^30^ Our results offer a fuller picture of how changes in cigarette consumption have varied according to the type of cigarettes smoked and across subgroups of smokers. There are three key findings.

First, remaining smokers are increasingly using hand-rolled tobacco over manufactured cigarettes. In 2008, the majority (69.4%) mainly or exclusively smoked manufactured cigarettes and, on average, twice as many manufactured cigarettes as hand-rolled cigarettes were consumed by smokers each day (9.5 vs. 4.2). By 2023, more than half (52.1%) mainly or exclusively smoked hand-rolled cigarettes and, on average, the number of manufactured and hand-rolled cigarettes consumed each day was similar (5.0 vs. 5.6). It is likely that many smokers will have switched from more expensive manufactured cigarettes to cheaper hand-rolled cigarettes^46^ to avoid spending more on tobacco as prices have risen.^43,46,47^ Young people may also increasingly prefer to start smoking with cheaper, hand-rolled cigarettes. Household budgets have been affected by a cost-of-living crisis England since late 2021^48^ which may have provided increased motivation to switch to (or take up smoking with) cheaper cigarettes in recent years. These results make a strong case for further higher relative tax increases on hand-rolled tobacco products to bring their cost more in line with manufactured cigarettes.

Second, the decline in overall cigarette consumption was non-linear and has stalled in recent years. While the average number of cigarettes consumed by smokers each day fell steadily between 2008 and 2019, there was virtually no change between late 2019 and 2023. The Covid-19 pandemic may have been an influencing factor: new working from home arrangements are generally more permissive of more regular smoking breaks, which may have disrupted the declining trend in daily cigarette consumption. Studies that looked at cigarette consumption during Covid lockdowns suggested that while some smokers reduced their consumption, others increased theirs,^49–52^ which may explain the overall stagnation. Reasons for increased consumption included opportunity, stress, and boredom.^49–51^ The number of manufactured cigarettes consumed has continued to decline, albeit more slowly than in earlier years, but this has been offset by a continued rise in hand-rolled cigarette consumption.

Finally, while significant changes in cigarette consumption were observed consistently (and in the same direction) across subgroups of smokers, the extent of these changes varied. Changes in the average number of any, manufactured, and hand-rolled cigarettes smoked per day were relatively more pronounced among non-daily and younger smokers. There was both a greater overall decline in cigarette consumption, and possibly a greater degree of switching from manufactured to hand-rolled cigarettes, among these groups. These groups are typically less dependent^53^ so it is possible they may be more willing to switch products in response to price increases or other economic pressures. Another potential explanation is that increasing use of e-cigarettes among young adults^54,55^ may have contributed to greater declines in cigarette consumption in this group. Consistent with this, we found patterns of cigarette consumption differed by vaping status. In 2008, dual users of cigarettes and e-cigarettes consumed a higher number of cigarettes per day than those who exclusively smoked, but this reversed over the study period such that by 2023, dual users reported lower consumption of cigarettes than smokers who did not vape. E-cigarettes appear to have initially attracted people who smoked more heavily^56^ but, over time, have either become increasingly popular among lighter smokers,^57^ helped people to reduce their cigarette consumption,^58,59^ or both.

These findings have implications for public health. All else being equal, reduced cigarette consumption will likely carry some health benefits at the population level. Even with compensatory changes in the way people smoke (i.e., smoking each cigarette harder to titrate their nicotine intake to the desired level),^12,13^ reducing the number of cigarettes they smoke each day will likely decrease the risk of disease at the individual level.^3–10^ Even relatively small improvements at the individual level can translate into noticeable reductions in smoking’s disease burden at a population level, with benefits for both smokers and those exposed to second-hand smoke. While average cigarette consumption has fallen over the past 15 years, this declining trend has stalled (and possibly reversed in some population groups). The availability of cheap, hand-rolled tobacco appears to be undermining policies that aim to reduce smoking by raising the price of tobacco (e.g., through taxation) and could be targeted to reignite the decline in cigarette consumption.

Strengths of this study include the large, nationally-representative sample and monthly data collection over a 15-year period. There were also limitations. First, the mode of data collection changed from face-to-face to telephone interviews in April 2020, when the Covid-19 pandemic started. Nonetheless, the two methods show good comparability^40–42^ and there is no obvious reason to expect participants to respond differently to the question assessing cigarette consumption over time. Secondly, e-cigarette use was not assessed before April 2011, but prevalence of use in England was very low before this,^60^ limiting scope to examine trends over a longer period if data had been available. In addition, the item assessing frequency of vaping did not distinguish between use of e-cigarettes specifically and use of other nicotine products. Although the majority (84.4%) of participants who were using e-cigarettes did not report use of any other nicotine products, it is possible that this variable did not accurately reflect vaping frequency among those who used both e-cigarettes and NRT, so these findings should be interpreted with some caution. Finally, as a household survey, the sample excluded people living in institutions or experiencing homelessness, so our findings may not be representative of changes in cigarette consumption among these groups.

In conclusion, the number of cigarettes smokers in England consumed each day has fallen by almost a quarter since 2008, but there has been little change since 2019. There has been a sharp decline in consumption of manufactured cigarettes and an increase in consumption of hand-rolled cigarettes, as smokers have increasingly shifted towards using hand-rolled tobacco.

## Declarations

## Funding

Cancer Research UK (PRCRPG-Nov21\100002) funded the Smoking Toolkit Study data collection and the salaries of Sarah Jackson and Harry Tattan-Birch. For the purpose of Open Access, the author has applied a CC BY public copyright licence to any Author Accepted Manuscript version arising from this submission.

## Competing interests

Jamie Brown has received unrestricted research funding from Pfizer and J&J, who manufacture smoking cessation medications. Lion Shahab has received honoraria for talks, an unrestricted research grant and travel expenses to attend meetings and workshops from Pfizer, and has acted as paid reviewer for grant awarding bodies and as a paid consultant for health care companies. All authors declare no financial links with tobacco companies, e-cigarette manufacturers, or their representatives.

## Author contributions

All authors contributed to the study conception and design. Data analysis was performed by Sarah Jackson. The first draft of the manuscript was written by Sarah Jackson and all authors commented on previous versions of the manuscript. All authors read and approved the final manuscript.

## Ethics approval

Ethical approval for the STS was granted originally by the UCL Ethics Committee (ID 0498/001). The data are not collected by UCL and are anonymized when received by UCL.

## Consent to participate

Informed consent was obtained from all individual participants included in the study.

## Data Availability

All data produced in the present study are available upon reasonable request to the authors.

## Notes

### Clinical Protocols

https://osf.io/bg7dv/

