## Supplementary material for "Trends in daily cigarette consumption among smokers: a population study in England, 2008-2023": Table S1

### Table S1. Sample characteristics (*n*=57,778)

| **Characteristic** | **Weighted proportion** |
| --- | --- |
| Age (years) |  |
| Mean (SD) | 41.7 (16.2) |
| 16-24 | 17.1% |
| 25-34 | 22.6% |
| 35-44 | 19.6% |
| 45-54 | 17.8% |
| 55-64 | 12.6% |
| ≥65 | 10.3% |
| Gender |  |
| Men | 52.4% |
| Women | 47.4% |
| Other | 0.2% |
| Missing, *n***^1^** | 25 |
| Occupational social grade |  |
| ABC1 (more advantaged) | 39.5% |
| C2DE (less advantaged) | 60.5% |
| Region in England |  |
| North | 32.0% |
| Midlands | 30.4% |
| South | 37.6% |
| Main type of cigarettes smoked |  |
| Manufactured | 57.4% |
| Hand-rolled | 42.6% |
| Missing, *n***^1^** | 955 |
| Frequency of smoking |  |
| Non-daily | 13.3% |
| Daily | 86.7% |
| Current use of NRT^2^ |  |
| No | 86.8% |
| Yes | 13.2% |
| Missing, *n***^1^** | 2,267 |
| Current vaping^2,3^ |  |
| No | 83.8% |
| Yes | 16.2% |
| Missing, *n***^1^** | 2,267 |
| Frequency of vaping^4^ |  |
| Non-daily | 36.6% |
| Daily | 63.4% |
| Missing, *n***^1^** | 1,247 |

Note: Data are shown as weighted column percentages, unless otherwise specified. There were some missing data (unweighted *n*s indicated in the table); valid percentages are shown for ease of interpretation.

^1^ Unweighted sample size.

^2^ Variable not assessed in certain waves (May/July/September/November 2012, January/March 2013). Missing cases include participants surveyed in these waves.

^3^ Among those surveyed from April 2011 onwards (unweighted *n*=48,427).

^4^ Among those reporting current vaping, April 2011 onwards (unweighted *n*=7,114). Variable not assessed in certain waves (May/June/August/September/November/December 2022, February/March/May/August/September 2023). Missing cases include participants surveyed in these waves.

### Table S2. Daily overall cigarette consumption by year among adult smokers in England, 2008-2023

|  | **Cigarettes per day, mean [95% CI]^1^** | | | | | | | | | | | | | | | |
| --- | --- | --- | --- | --- | --- | --- | --- | --- | --- | --- | --- | --- | --- | --- | --- | --- |
|  | **2008** | **2009** | **2010** | **2011** | **2012** | **2013** | **2014** | **2015** | **2016** | **2017** | **2018** | **2019** | **2020** | **2021** | **2022** | **2023^2^** |
| All adult (≥18y) smokers | 13.5  [13.2–13.7] | 13.1  [12.9–13.4] | 12.8  [12.6–13.0] | 12.4  [12.2–12.7] | 12.2  [11.9–12.5] | 11.8  [11.6–12.1] | 11.4  [11.2–11.7] | 11.3  [11.1–11.6] | 11.3  [11.0–11.5] | 10.9  [10.6–11.2] | 10.6  [10.3–10.9] | 10.4  [10.1–10.7] | 10.8  [10.4–11.2] | 10.5  [10.1–10.8] | 10.3  [9.9–10.7] | 11.0  [10.5–11.4] |
| Main type of cigarettes smoked |  |  |  |  |  |  |  |  |  |  |  |  |  |  |  |  |
| Manufactured | 13.2  [12.9–13.5] | 12.8  [12.5–13.1] | 12.3  [12.0–12.6] | 11.9  [11.6–12.2] | 11.7  [11.4–12.1] | 11.2  [10.8–11.5] | 11.1  [10.7–11.5] | 10.6  [10.3–11.0] | 11.2  [10.8–11.5] | 10.7  [10.3–11.1] | 10.2  [9.8–10.6] | 10.4  [10.0–10.9] | 10.5  [10.0–11.0] | 10.1  [9.7–10.6] | 10.5  [9.9–11.1] | 11.0  [10.4–11.7] |
| Hand-rolled | 14.1  [13.6–14.6] | 13.8  [13.3–14.2] | 13.7  [13.3–14.1] | 13.4  [13.0–13.8] | 12.9  [12.5–13.4] | 12.6  [12.2–13] | 12.1  [11.6–12.5] | 12.2  [11.7–12.6] | 11.4  [11.0–11.9] | 11.1  [10.7–11.6] | 11.0  [10.6–11.5] | 10.5  [10.1–10.9] | 11.0  [10.5–11.6] | 10.9  [10.4–11.4] | 10.1  [9.5–10.6] | 11.1  [10.4–11.7] |
| Frequency of smoking |  |  |  |  |  |  |  |  |  |  |  |  |  |  |  |  |
| Non-daily | 5.1  [4.5–5.7] | 5.1  [4.6–5.7] | 5.5  [4.9–6.0] | 5.0  [4.4–5.5] | 5.1  [4.3–5.9] | 4.7  [4.2–5.3] | 3.8  [3.2–4.3] | 3.6  [3.1–4.1] | 3.9  [3.3–4.5] | 3.4  [3.0–3.9] | 3.3  [2.9–3.7] | 3.1  [2.7–3.5] | 3.0  [2.7–3.4] | 3.6  [3.1–4.0] | 3.1  [2.8–3.5] | 3.0  [2.6–3.3] |
| Daily | 14.3  [14.1–14.6] | 14.2  [14.0–14.5] | 13.7  [13.5–14.0] | 13.1  [12.9–13.4] | 13.0  [12.7–13.2] | 12.8  [12.5–13.1] | 12.4  [12.1–12.7] | 12.4  [12.1–12.7] | 12.2  [11.9–12.5] | 12.1  [11.7–12.4] | 11.9  [11.6–12.2] | 11.5  [11.2–11.8] | 12.7  [12.3–13.1] | 12.4  [12.0–12.8] | 12.4  [11.9–12.8] | 13.3  [12.8–13.8] |
| Age (years) |  |  |  |  |  |  |  |  |  |  |  |  |  |  |  |  |
| 18-24 | 12.0  [11.4–12.6] | 10.9  [10.4–11.4] | 10.7  [10.2–11.2] | 10.3  [9.8–10.9] | 10.1  [9.5–10.8] | 10.0  [9.4–10.5] | 9.4  [8.9–10.0] | 9.6  [9.1–10.1] | 9.1  [8.5–9.7] | 8.4  [7.8–9.0] | 7.7  [7.2–8.3] | 8.0  [7.4–8.7] | 8.7  [7.9–9.5] | 8.0  [7.1–8.9] | 7.6  [6.7–8.5] | 6.7  [5.8–7.6] |
| 25-34 | 11.7  [11.1–12.3] | 11.4  [10.9–11.9] | 10.9  [10.5–11.4] | 11.0  [10.5–11.4] | 10.8  [10.3–11.3] | 10.3  [9.8–10.8] | 10.0  [9.5–10.5] | 9.9  [9.4–10.5] | 10.1  [9.6–10.7] | 9.8  [9.3–10.4] | 9.4  [8.8–10.0] | 9.7  [9.1–10.3] | 10.3  [9.5–11.0] | 9.1  [8.5–9.8] | 9.4  [8.5–10.3] | 9.8  [8.6–10.9] |
| 35-44 | 14.1  [13.5–14.7] | 13.3  [12.8–13.8] | 13.2  [12.7–13.8] | 12.8  [12.2–13.4] | 12.2  [11.6–12.9] | 12.1  [11.6–12.7] | 11.5  [10.8–12.2] | 11.3  [10.6–12.0] | 11.3  [10.6–11.9] | 11.2  [10.5–12.0] | 10.7  [10.1–11.4] | 9.7  [9.1–10.4] | 11.1  [10.0–12.1] | 9.8  [8.9–10.6] | 10.3  [9.5–11.1] | 10.9  [9.8–12.0] |
| 45-54 | 15.6  [14.9–16.3] | 15.2  [14.6–15.9] | 14.6  [14.1–15.2] | 14.0  [13.3–14.7] | 14.6  [13.8–15.3] | 13.2  [12.5–13.9] | 13.2  [12.5–13.9] | 12.9  [12.1–13.7] | 12.5  [11.8–13.2] | 12.1  [11.3–12.9] | 12.1  [11.3–12.9] | 12.1  [11.3–12.8] | 12.0  [10.9–13.1] | 12.4  [11.5–13.2] | 12.1  [11.2–13.1] | 12.2  [11.2–13.2] |
| 55-64 | 15.3  [14.5–16.1] | 16.0  [15.3–16.8] | 14.9  [14.2–15.7] | 14.0  [13.3–14.8] | 13.6  [12.8–14.3] | 14.2  [13.4–15.0] | 13.0  [12.2–13.8] | 13.1  [12.4–13.8] | 14.0  [13.1–14.9] | 12.5  [11.7–13.2] | 13.2  [12.4–14.0] | 12.3  [11.5–13.2] | 12.6  [11.7–13.5] | 13.3  [12.3–14.2] | 12.3  [11.2–13.4] | 13.1  [12.0–14.1] |
| ≥65 | 12.9  [12.0–13.7] | 13.5  [12.7–14.4] | 13.7  [12.9–14.4] | 13.1  [12.3–13.8] | 12.3  [11.6–13.0] | 12.3  [11.5–13.0] | 12.8  [12.0–13.5] | 12.6  [11.8–13.5] | 11.6  [10.8–12.4] | 12.7  [11.9–13.6] | 11.8  [11.1–12.5] | 11.5  [10.8–12.2] | 10.8  [10.1–11.6] | 12.6  [11.8–13.4] | 11.3  [10.4–12.2] | 13.8  [12.6–14.9] |
| Gender |  |  |  |  |  |  |  |  |  |  |  |  |  |  |  |  |
| Men | 14.3  [13.9–14.8] | 13.8  [13.5–14.2] | 13.5  [13.2–13.9] | 13.2  [12.8–13.6] | 12.9  [12.5–13.3] | 12.4  [12.0–12.8] | 12.0  [11.6–12.4] | 11.7  [11.3–12.1] | 11.9  [11.4–12.3] | 11.4  [10.9–11.8] | 11.3  [10.8–11.7] | 10.8  [10.4–11.2] | 11.5  [10.9–12.0] | 11.4  [10.9–12.0] | 11.3  [10.7–11.9] | 11.5  [10.8–12.2] |
| Women | 12.6  [12.2–12.9] | 12.4  [12.1–12.7] | 11.9  [11.7–12.2] | 11.6  [11.2–11.9] | 11.4  [11.0–11.7] | 11.1  [10.8–11.4] | 10.8  [10.4–11.2] | 11.0  [10.6–11.4] | 10.6  [10.2–10.9] | 10.4  [10.0–10.8] | 9.8  [9.4–10.1] | 10.0  [9.6–10.4] | 10.0  [9.5–10.5] | 9.5  [9.1–9.9] | 9.0  [8.5–9.4] | 10.2  [9.6–10.7] |
| Occupational social grade |  |  |  |  |  |  |  |  |  |  |  |  |  |  |  |  |
| ABC1 (more advantaged) | 12.2  [11.7–12.6] | 12.2  [11.8–12.6] | 11.6  [11.2–12.0] | 11.5  [11.0–11.9] | 11.5  [11.0–11.9] | 10.4  [10.0–10.8] | 10.4  [9.9–10.8] | 9.9  [9.5–10.4] | 9.5  [9.1–9.9] | 9.3  [8.9–9.7] | 9.1  [8.7–9.5] | 9.0  [8.6–9.4] | 9.4  [9.0–9.9] | 9.6  [9.1–10.0] | 9.5  [8.9–10.0] | 9.8  [9.2–10.4] |
| C2DE (less advantaged) | 14.3  [13.9–14.6] | 13.8  [13.5–14.1] | 13.7  [13.4–14.0] | 13.1  [12.8–13.4] | 12.7  [12.4–13.0] | 12.7  [12.3–13.0] | 12.1  [11.7–12.4] | 12.2  [11.8–12.5] | 12.4  [12.0–12.8] | 11.9  [11.5–12.3] | 11.5  [11.1–11.9] | 11.3  [10.9–11.7] | 11.6  [11.1–12.1] | 11.2  [10.7–11.7] | 10.9  [10.4–11.4] | 11.8  [11.2–12.5] |
| Region in England |  |  |  |  |  |  |  |  |  |  |  |  |  |  |  |  |
| North | 14.2  [13.8–14.7] | 13.7  [13.2–14.1] | 14.0  [13.6–14.4] | 13.4  [12.9–13.9] | 13.0  [12.5–13.5] | 12.4  [12.0–12.8] | 12.0  [11.5–12.4] | 12.4  [12.0–12.9] | 12.0  [11.5–12.5] | 11.7  [11.1–12.2] | 11.0  [10.5–11.5] | 11.1  [10.6–11.6] | 11.4  [10.7–12.1] | 11.3  [10.7–12] | 10.9  [10.2–11.6] | 11.8  [10.9–12.7] |
| Midlands | 13.5  [13.0–14.0] | 13.6  [13.1–14.0] | 12.6  [12.2–13.1] | 12.7  [12.3–13.2] | 12.2  [11.7–12.7] | 12.2  [11.6–12.7] | 11.2  [10.7–11.7] | 11.2  [10.6–11.7] | 11.4  [10.9–11.9] | 11.3  [10.7–11.8] | 11.1  [10.6–11.6] | 11.0  [10.4–11.6] | 11.1  [10.4–11.7] | 10.5  [9.9–11.1] | 10.6  [10.0–11.3] | 11.9  [11.0–12.8] |
| South | 12.6  [12.1–13.1] | 12.3  [11.9–12.7] | 11.8  [11.4–12.2] | 11.3  [10.9–11.7] | 11.5  [11.1–12.0] | 11.0  [10.6–11.4] | 11.1  [10.7–11.6] | 10.5  [10.0–11.0] | 10.5  [10.1–11.0] | 10.0  [9.5–10.4] | 9.8  [9.3–10.3] | 9.5  [9.0–9.9] | 10.1  [9.5–10.7] | 9.9  [9.3–10.4] | 9.7  [9.0–10.3] | 9.9  [9.2–10.5] |
| Current use of NRT |  |  |  |  |  |  |  |  |  |  |  |  |  |  |  |  |
| No | 13.3  [13.0–13.6] | 12.9  [12.7–13.2] | 12.8  [12.5–13.0] | 12.3  [12.0–12.6] | 12.3  [11.9–12.7] | 11.8  [11.5–12.1] | 11.4  [11.1–11.7] | 11.3  [11.0–11.6] | 11.3  [11.0–11.6] | 10.9  [10.6–11.2] | 10.4  [10.1–10.7] | 10.5  [10.2–10.8] | 10.6  [10.2–11.0] | 10.4  [10.0–10.7] | 10.2  [9.8–10.6] | 10.8  [10.3–11.2] |
| Yes | 14.3  [13.7–15.0] | 13.9  [13.4–14.5] | 12.8  [12.3–13.4] | 12.9  [12.3–13.6] | 12.6  [11.6–13.6] | 12.7  [11.9–13.5] | 11.5  [10.8–12.3] | 12.1  [11.1–13.0] | 11.0  [10.1–11.8] | 11.1  [10.2–11.9] | 11.7  [10.8–12.6] | 10.2  [9.3–11.2] | 12.1  [11.1–13.2] | 11.4  [10.3–12.5] | 11.2  [10.0–12.4] | 12.5  [10.8–14.2] |
| Current vaping^3^ |  |  |  |  |  |  |  |  |  |  |  |  |  |  |  |  |
| No | - | - | - | 12.4  [12.1–12.6] | 12.3  [11.9–12.7] | 11.7  [11.4–12.1] | 11.4  [11.1–11.7] | 11.2  [10.8–11.5] | 11.3  [11.0–11.6] | 10.8  [10.5–11.2] | 10.6  [10.3–10.9] | 10.6  [10.3–10.9] | 10.8  [10.4–11.3] | 10.6  [10.2–11.0] | 10.5  [10.0–10.9] | 11.6  [11.0–12.1] |
| Yes | - | - | - | 14.5  [12.7–16.2] | 12.8  [11.7–13.9] | 12.6  [11.9–13.3] | 11.7  [11.1–12.3] | 12.0  [11.4–12.7] | 11.0  [10.4–11.6] | 11.1  [10.4–11.7] | 10.4  [9.8–11.0] | 9.7  [9.0–10.3] | 10.6  [9.8–11.4] | 10.0  [9.3–10.6] | 9.9  [9.1–10.6] | 9.7  [8.9–10.5] |
| Non-daily | - | - | - | 15.4  [12.5–18.4] | 14.2  [12.1–16.4] | 12.5  [11.5–13.6] | 12.2  [11.2–13.3] | 12.4  [11.5–13.3] | 11.6  [10.6–12.6] | 11.3  [10.3–12.3] | 10.9  [9.8–11.9] | 10.2  [8.8–11.7] | 11.4  [9.8–13.0] | 11.1  [9.6–12.6] | 9.6  [7.6–11.7] | 13.5  [11.3–15.6] |
| Daily | - | - | - | 13.9  [11.7–16.1] | 12  [10.8–13.2] | 12.7  [11.7–13.8] | 11.3  [10.6–12.1] | 11.4  [10.6–12.2] | 10.6  [9.8–11.4] | 10.9  [10.1–11.8] | 10.3  [9.5–11.1] | 9.3  [8.6–10.1] | 10.2  [9.2–11.2] | 9.4  [8.6–10.3] | 10.8  [8.9–12.6] | 8.5  [7.2–9.7] |

^1^ Data are weighted means.

**^2^** Data for 2023 are based on January-September only.

**^3^** Vaping status has only been assessed since April 2011, so no data are available for 2008-2010 and data for 2011 are based on April-December only.

### Table S3. Daily manufactured cigarette consumption by year among adult smokers in England, 2008-2023

|  | **Manufactured cigarettes per day, mean [95% CI]^1^** | | | | | | | | | | | | | | | |
| --- | --- | --- | --- | --- | --- | --- | --- | --- | --- | --- | --- | --- | --- | --- | --- | --- |
|  | **2008** | **2009** | **2010** | **2011** | **2012** | **2013** | **2014** | **2015** | **2016** | **2017** | **2018** | **2019** | **2020** | **2021** | **2022** | **2023^2^** |
| All adult (≥18y) smokers | 9.3  [9.0–9.6] | 8.7  [8.4–8.9] | 8.1  [7.9–8.4] | 7.7  [7.4–8.0] | 7.3  [7.0–7.5] | 6.3  [6.0–6.6] | 6.4  [6.1–6.7] | 5.9  [5.6–6.2] | 6.0  [5.8–6.3] | 5.5  [5.3–5.8] | 5.3  [5.0–5.6] | 5.4  [5.1–5.7] | 5.4  [5.0–5.7] | 5.0  [4.7–5.3] | 4.9  [4.6–5.3] | 5.4  [5.0–5.8] |
| Main type of cigarettes smoked |  |  |  |  |  |  |  |  |  |  |  |  |  |  |  |  |
| Manufactured | 13.0  [12.6–13.3] | 12.6  [12.3–12.9] | 12.1  [11.8–12.4] | 11.7  [11.4–12.0] | 11.6  [11.2–11.9] | 11.0  [10.7–11.3] | 10.9  [10.6–11.3] | 10.5  [10.1–10.8] | 11.1  [10.7–11.4] | 10.6  [10.2–11] | 10.0  [9.6–10.4] | 10.3  [9.9–10.8] | 10.4  [9.9–10.9] | 10.0  [9.5–10.4] | 10.2  [9.7–10.8] | 10.9  [10.2–11.5] |
| Hand-rolled | 0.9  [0.7–1.0] | 0.8  [0.6–0.9] | 0.6  [0.5–0.7] | 0.5  [0.4–0.6] | 0.5  [0.4–0.6] | 0.5  [0.4–0.6] | 0.5  [0.4–0.6] | 0.4  [0.3–0.4] | 0.3  [0.2–0.4] | 0.2  [0.2–0.3] | 0.2  [0.2–0.3] | 0.3  [0.2–0.3] | 0.3  [0.2–0.4] | 0.3  [0.2–0.3] | 0.4  [0.2–0.5] | 0.3  [0.2–0.4] |
| Frequency of smoking |  |  |  |  |  |  |  |  |  |  |  |  |  |  |  |  |
| Non-daily | 3.9  [3.4–4.5] | 3.9  [3.5–4.4] | 4.3  [3.7–4.8] | 3.6  [3.1–4.1] | 3.6  [2.9–4.3] | 3.1  [2.6–3.5] | 2.4  [2.0–2.9] | 2.4  [2.0–2.9] | 2.4  [2.0–2.9] | 2.1  [1.7–2.5] | 2.0  [1.6–2.4] | 2.1  [1.7–2.5] | 1.8  [1.4–2.1] | 2.2  [1.8–2.6] | 1.7  [1.4–1.9] | 1.8  [1.5–2.1] |
| Daily | 9.8  [9.5–10.1] | 9.3  [9.0–9.6] | 8.6  [8.4–8.9] | 8.1  [7.8–8.4] | 7.7  [7.4–8.0] | 6.8  [6.5–7.0] | 6.9  [6.6–7.2] | 6.4  [6.1–6.7] | 6.5  [6.2–6.8] | 6.1  [5.8–6.4] | 5.9  [5.6–6.2] | 5.9  [5.5–6.2] | 6.3  [5.9–6.7] | 5.8  [5.4–6.2] | 5.8  [5.4–6.3] | 6.4  [5.9–6.9] |
| Age (years) |  |  |  |  |  |  |  |  |  |  |  |  |  |  |  |  |
| 18-24 | 8.4  [7.8–9.0] | 7.1  [6.6–7.7] | 6.6  [6.1–7.1] | 6.4  [5.8–6.9] | 5.5  [5.0–6.1] | 4.5  [4.0–5.0] | 4.5  [3.9–5.0] | 4.2  [3.7–4.7] | 3.8  [3.3–4.3] | 3.2  [2.7–3.7] | 2.8  [2.4–3.2] | 2.9  [2.3–3.4] | 3.0  [2.4–3.6] | 2.6  [2.1–3.2] | 3.3  [2.5–4.0] | 2.1  [1.5–2.7] |
| 25-34 | 8.1  [7.5–8.7] | 7.4  [6.8–7.9] | 7.2  [6.7–7.7] | 7.0  [6.5–7.5] | 6.3  [5.8–6.7] | 5.5  [5.0–6.0] | 5.4  [4.9–6.0] | 5.1  [4.6–5.6] | 5.4  [4.8–6.0] | 5.1  [4.5–5.7] | 4.8  [4.3–5.4] | 4.6  [4.0–5.3] | 4.4  [3.7–5.1] | 3.3  [2.8–3.8] | 3.4  [2.8–4.1] | 3.4  [2.7–4.2] |
| 35-44 | 9.4  [8.7–10.0] | 8.0  [7.4–8.5] | 7.7  [7.2–8.2] | 7.7  [7.1–8.3] | 7.3  [6.7–8.0] | 6.6  [6.0–7.2] | 6.4  [5.7–7.1] | 6.0  [5.4–6.6] | 6.1  [5.4–6.8] | 5.1  [4.5–5.8] | 5.1  [4.5–5.8] | 5.2  [4.6–5.8] | 5.4  [4.6–6.2] | 4.6  [4.0–5.3] | 4.7  [3.9–5.5] | 4.8  [3.7–6.0] |
| 45-54 | 9.9  [9.1–10.6] | 10.1  [9.4–10.8] | 9.0  [8.3–9.6] | 7.9  [7.3–8.6] | 8.1  [7.3–8.9] | 6.7  [6.1–7.4] | 7.4  [6.7–8.2] | 6.4  [5.6–7.1] | 6.9  [6.2–7.6] | 6.3  [5.6–7.0] | 6.0  [5.3–6.7] | 5.8  [5.0–6.6] | 6.0  [5.1–6.9] | 6.6  [5.8–7.4] | 6.1  [5.2–7.1] | 5.9  [5.0–6.8] |
| 55-64 | 10.7  [9.8–11.6] | 11.1  [10.3–12.0] | 10.0  [9.2–10.7] | 8.9  [8.1–9.7] | 8.1  [7.3–9.0] | 8.2  [7.4–9.0] | 7.2  [6.4–8.0] | 7.4  [6.6–8.2] | 7.4  [6.4–8.3] | 6.7  [5.9–7.4] | 6.9  [6.0–7.7] | 7.0  [6.1–7.9] | 7.7  [6.7–8.6] | 7.3  [6.3–8.3] | 6.5  [5.4–7.5] | 7.1  [6.1–8.2] |
| ≥65 | 10.8  [9.9–11.7] | 10.5  [9.6–11.4] | 10.2  [9.4–11.0] | 9.4  [8.7–10.2] | 9.3  [8.5–10.1] | 7.6  [6.9–8.3] | 9.0  [8.2–9.8] | 7.9  [7.0–8.7] | 7.7  [7.0–8.4] | 8.6  [7.6–9.5] | 7.6  [6.7–8.4] | 8.1  [7.2–8.9] | 7.9  [7.0–8.7] | 8.3  [7.4–9.3] | 7.3  [6.3–8.4] | 9.8  [8.6–11.0] |
| Gender |  |  |  |  |  |  |  |  |  |  |  |  |  |  |  |  |
| Men | 9.0  [8.5–9.4] | 8.0  [7.6–8.4] | 7.6  [7.2–7.9] | 7.3  [6.9–7.7] | 6.8  [6.4–7.2] | 5.7  [5.4–6.1] | 6.0  [5.6–6.4] | 5.6  [5.2–6.0] | 6.0  [5.6–6.4] | 5.2  [4.8–5.6] | 5.2  [4.8–5.6] | 5.3  [4.9–5.7] | 5.5  [5.0–6.0] | 5.3  [4.8–5.7] | 5.3  [4.7–5.8] | 5.2  [4.6–5.8] |
| Women | 9.6  [9.2–9.9] | 9.3  [9.0–9.7] | 8.8  [8.5–9.1] | 8.2  [7.8–8.5] | 7.8  [7.5–8.2] | 7.0  [6.6–7.3] | 6.8  [6.5–7.2] | 6.2  [5.8–6.6] | 6.1  [5.7–6.4] | 5.9  [5.5–6.3] | 5.5  [5.1–5.8] | 5.5  [5.1–5.9] | 5.3  [4.9–5.7] | 4.8  [4.5–5.2] | 4.3  [3.9–4.8] | 5.4  [4.9–5.9] |
| Occupational social grade |  |  |  |  |  |  |  |  |  |  |  |  |  |  |  |  |
| ABC1 (more advantaged) | 9.5  [9.0–10.0] | 9.0  [8.5–9.4] | 8.3  [7.9–8.7] | 8.2  [7.8–8.7] | 7.8  [7.3–8.3] | 6.4  [6.0–6.8] | 6.7  [6.2–7.1] | 6.1  [5.6–6.5] | 6.1  [5.7–6.5] | 5.6  [5.2–6.0] | 5.2  [4.8–5.5] | 5.0  [4.6–5.5] | 5.2  [4.8–5.7] | 5.4  [5.0–5.8] | 5.3  [4.9–5.8] | 5.4  [4.9–6.0] |
| C2DE (less advantaged) | 9.1  [8.8–9.5] | 8.4  [8.1–8.8] | 8.0  [7.7–8.3] | 7.3  [7.0–7.7] | 6.9  [6.6–7.3] | 6.3  [5.9–6.6] | 6.2  [5.9–6.6] | 5.8  [5.5–6.1] | 6.0  [5.7–6.4] | 5.5  [5.1–5.9] | 5.4  [5.0–5.8] | 5.6  [5.2–6.0] | 5.4  [5.0–5.9] | 4.8  [4.4–5.2] | 4.6  [4.1–5.1] | 5.3  [4.8–5.9] |
| Region in England |  |  |  |  |  |  |  |  |  |  |  |  |  |  |  |  |
| North | 10.4  [9.9–10.9] | 9.7  [9.2–10.2] | 9.6  [9.2–10.1] | 9.1  [8.7–9.6] | 8.3  [7.8–8.8] | 6.9  [6.5–7.3] | 6.5  [6.0–7.0] | 6.4  [6.0–6.9] | 6.5  [6.0–7.0] | 6.1  [5.5–6.6] | 5.8  [5.4–6.3] | 5.8  [5.2–6.3] | 5.8  [5.2–6.4] | 5.7  [5.1–6.3] | 5.5  [4.8–6.2] | 6.1  [5.4–6.9] |
| Midlands | 8.8  [8.2–9.3] | 8.5  [8.0–9.0] | 7.7  [7.2–8.1] | 7.7  [7.2–8.2] | 7.0  [6.5–7.4] | 6.0  [5.5–6.4] | 6.7  [6.2–7.2] | 5.9  [5.4–6.4] | 6.4  [5.8–6.9] | 5.9  [5.4–6.4] | 5.3  [4.8–5.8] | 5.6  [5.1–6.2] | 5.3  [4.7–5.9] | 4.9  [4.4–5.5] | 4.9  [4.3–5.6] | 5.5  [4.7–6.3] |
| South | 8.5  [8.0–9.0] | 7.9  [7.4–8.3] | 7.2  [6.8–7.6] | 6.5  [6.1–6.9] | 6.6  [6.2–7.1] | 6.0  [5.6–6.4] | 6.1  [5.6–6.6] | 5.4  [4.9–5.9] | 5.4  [5.0–5.8] | 4.9  [4.4–5.3] | 4.9  [4.5–5.4] | 4.9  [4.4–5.3] | 5.1  [4.6–5.7] | 4.6  [4.2–5.1] | 4.4  [3.9–4.9] | 4.8  [4.2–5.4] |
| Current use of NRT |  |  |  |  |  |  |  |  |  |  |  |  |  |  |  |  |
| No | 9.0  [8.7–9.3] | 8.3  [8.0–8.6] | 8.0  [7.7–8.2] | 7.4  [7.2–7.7] | 7.4  [7.0–7.7] | 6.1  [5.8–6.4] | 6.3  [6.0–6.6] | 5.9  [5.6–6.2] | 6.0  [5.7–6.3] | 5.5  [5.2–5.8] | 5.2  [4.9–5.5] | 5.4  [5.1–5.7] | 5.2  [4.9–5.6] | 5.0  [4.7–5.3] | 4.8  [4.4–5.2] | 5.3  [4.9–5.8] |
| Yes | 10.7  [9.9–11.4] | 9.9  [9.3–10.5] | 8.9  [8.3–9.5] | 9.0  [8.3–9.7] | 8.3  [7.3–9.4] | 7.1  [6.3–7.8] | 7.3  [6.5–8.1] | 6.0  [5.1–6.9] | 6.2  [5.3–7.1] | 6.4  [5.5–7.3] | 6.4  [5.6–7.3] | 5.1  [4.3–6.0] | 6.6  [5.6–7.7] | 5.3  [4.6–6.1] | 5.6  [4.6–6.7] | 5.7  [4.7–6.8] |
| Current vaping^3^ |  |  |  |  |  |  |  |  |  |  |  |  |  |  |  |  |
| No | - | - | - | 7.6  [7.3–7.9] | 7.4  [7.0–7.7] | 6.1  [5.8–6.4] | 6.3  [6.0–6.6] | 5.7  [5.4–6.0] | 6.1  [5.8–6.5] | 5.6  [5.3–5.9] | 5.3  [5.0–5.6] | 5.4  [5.1–5.8] | 5.3  [5.0–5.7] | 5.1  [4.7–5.4] | 4.9  [4.5–5.3] | 5.9  [5.4–6.4] |
| Yes | - | - | - | 12.1  [10.3–14.0] | 9.1  [7.9–10.4] | 7.2  [6.5–7.9] | 6.8  [6.2–7.4] | 6.6  [6.0–7.3] | 5.7  [5.0–6.3] | 5.4  [4.8–6.0] | 5.4  [4.8–6.1] | 5.2  [4.5–5.8] | 5.5  [4.7–6.2] | 4.9  [4.3–5.5] | 4.9  [4.2–5.6] | 4.2  [3.6–4.8] |
| Non-daily | - | - | - | 13.9  [10.8–17.0] | 8.7  [6.1–11.2] | 7.2  [6.2–8.3] | 6.9  [5.8–8.0] | 6.5  [5.5–7.5] | 6.2  [5.2–7.3] | 5.7  [4.7–6.7] | 5.9  [4.9–7.0] | 5.4  [4.1–6.7] | 5.8  [4.1–7.4] | 5.1  [3.8–6.4] | 3.7  [2.2–5.3] | 6.3  [4.0–8.6] |
| Daily | - | - | - | 11.1  [8.8–13.4] | 9.4  [8.0–10.7] | 7.6  [6.6–8.5] | 6.7  [6.0–7.4] | 6.4  [5.6–7.2] | 5.5  [4.7–6.3] | 5.3  [4.4–6.1] | 5.2  [4.4–6.1] | 5.1  [4.3–5.9] | 5.2  [4.3–6.2] | 4.9  [4.1–5.7] | 5.5  [4.1–6.9] | 3.7  [2.7–4.6] |

^1^ Data are weighted means.

**^2^** Data for 2023 are based on January-September only.

**^3^** Vaping status has only been assessed since April 2011, so no data are available for 2008-2010 and data for 2011 are based on April-December only.

### Table S4. Daily hand-rolled cigarette consumption by year among adult smokers in England, 2008-2023

|  | **Hand-rolled cigarettes per day, mean [95% CI]^1^** | | | | | | | | | | | | | | | |
| --- | --- | --- | --- | --- | --- | --- | --- | --- | --- | --- | --- | --- | --- | --- | --- | --- |
|  | **2008** | **2009** | **2010** | **2011** | **2012** | **2013** | **2014** | **2015** | **2016** | **2017** | **2018** | **2019** | **2020** | **2021** | **2022** | **2023^2^** |
| All adult (≥18y) smokers | 4.2  [4.0–4.4] | 4.5  [4.3–4.7] | 4.6  [4.4–4.9] | 4.7  [4.5–5.0] | 4.9  [4.7–5.2] | 5.5  [5.2–5.8] | 5.1  [4.8–5.4] | 5.4  [5.1–5.7] | 5.3  [5.0–5.5] | 5.4  [5.1–5.7] | 5.3  [5.0–5.6] | 5.1  [4.8–5.3] | 5.4  [5.0–5.8] | 5.5  [5.1–5.8] | 5.4  [5.0–5.7] | 5.7  [5.3–6.1] |
| Main type of cigarettes smoked |  |  |  |  |  |  |  |  |  |  |  |  |  |  |  |  |
| Manufactured | 0.2  [0.2–0.3] | 0.2  [0.2–0.2] | 0.2  [0.2–0.3] | 0.2  [0.1–0.2] | 0.2  [0.1–0.2] | 0.2  [0.2–0.3] | 0.2  [0.1–0.2] | 0.2  [0.1–0.2] | 0.1  [0.1–0.1] | 0.2  [0.1–0.2] | 0.1  [0.1–0.2] | 0.1  [0.1–0.2] | 0.1  [0.1–0.2] | 0.1  [0.1–0.2] | 0.3  [0.2–0.4] | 0.2  [0.1–0.2] |
| Hand-rolled | 13.2  [12.8–13.7] | 13.0  [12.6–13.4] | 13.1  [12.7–13.5] | 12.9  [12.5–13.4] | 12.4  [12.0–12.8] | 12.1  [11.7–12.5] | 11.6  [11.2–12.0] | 11.8  [11.4–12.3] | 11.1  [10.7–11.6] | 10.9  [10.5–11.3] | 10.8  [10.4–11.2] | 10.2  [9.8–10.6] | 10.7  [10.2–11.3] | 10.6  [10.1–11.1] | 9.7  [9.3–10.2] | 10.8  [10.1–11.4] |
| Frequency of smoking |  |  |  |  |  |  |  |  |  |  |  |  |  |  |  |  |
| Non-daily | 1.2  [0.8–1.5] | 1.2  [0.9–1.5] | 1.2  [0.9–1.5] | 1.4  [1.0–1.7] | 1.5  [1.1–2.0] | 1.6  [1.3–2.0] | 1.3  [1.0–1.6] | 1.2  [0.9–1.5] | 1.5  [1.0–2.0] | 1.3  [0.9–1.6] | 1.3  [1.0–1.5] | 1  [0.8–1.3] | 1.3  [1.0–1.5] | 1.3  [0.9–1.6] | 1.5  [1.2–1.8] | 1.2  [0.9–1.4] |
| Daily | 4.5  [4.3–4.8] | 4.9  [4.7–5.2] | 5.1  [4.8–5.3] | 5.0  [4.8–5.3] | 5.3  [5.0–5.6] | 6.0  [5.7–6.3] | 5.6  [5.3–5.9] | 6.0  [5.7–6.3] | 5.7  [5.4–6.0] | 6.0  [5.7–6.3] | 6.0  [5.7–6.3] | 5.6  [5.3–5.9] | 6.4  [6.0–6.9] | 6.6  [6.2–7.0] | 6.5  [6.1–6.9] | 7.0  [6.5–7.5] |
| Age (years) |  |  |  |  |  |  |  |  |  |  |  |  |  |  |  |  |
| 18-24 | 3.6  [3.0–4.1] | 3.8  [3.3–4.3] | 4.1  [3.6–4.6] | 4.0  [3.5–4.5] | 4.6  [4.0–5.2] | 5.5  [4.9–6.0] | 5.0  [4.5–5.5] | 5.3  [4.8–5.9] | 5.3  [4.7–6.0] | 5.2  [4.6–5.8] | 4.9  [4.3–5.4] | 5.2  [4.5–5.8] | 5.7  [4.9–6.5] | 5.3  [4.5–6.2] | 4.2  [3.6–4.9] | 4.6  [3.8–5.4] |
| 25-34 | 3.7  [3.2–4.1] | 4.0  [3.6–4.5] | 3.7  [3.3–4.1] | 4.0  [3.5–4.4] | 4.5  [4.1–5.0] | 4.7  [4.2–5.2] | 4.6  [4.1–5.1] | 4.9  [4.3–5.4] | 4.7  [4.1–5.3] | 4.8  [4.3–5.4] | 4.7  [4.1–5.3] | 5.1  [4.5–5.6] | 5.9  [5.2–6.7] | 5.8  [5.1–6.5] | 6.1  [5.2–6.9] | 6.4  [5.3–7.5] |
| 35-44 | 4.7  [4.2–5.3] | 5.4  [4.8–5.9] | 5.5  [5.0–6.0] | 5.1  [4.5–5.6] | 4.9  [4.3–5.5] | 5.5  [5.0–6.1] | 5.2  [4.6–5.9] | 5.3  [4.6–6.0] | 5.2  [4.5–5.9] | 6.1  [5.3–6.9] | 5.6  [4.9–6.2] | 4.6  [3.9–5.2] | 5.6  [4.6–6.6] | 5.1  [4.3–6.0] | 5.6  [4.8–6.3] | 6.1  [5.1–7.1] |
| 45-54 | 5.7  [5.1–6.4] | 5.2  [4.6–5.7] | 5.7  [5.1–6.2] | 6.1  [5.4–6.8] | 6.4  [5.7–7.2] | 6.6  [5.8–7.3] | 5.9  [5.2–6.6] | 6.5  [5.6–7.3] | 5.7  [5.0–6.4] | 5.9  [5.1–6.7] | 6.2  [5.4–7.0] | 6.1  [5.4–6.8] | 5.8  [4.9–6.8] | 5.9  [5.0–6.7] | 5.9  [5.1–6.7] | 6.3  [5.2–7.4] |
| 55-64 | 4.6  [3.8–5.4] | 4.9  [4.2–5.7] | 5.0  [4.2–5.7] | 5.1  [4.5–5.8] | 5.5  [4.7–6.2] | 6.0  [5.2–6.8] | 5.8  [5.0–6.6] | 5.8  [5.0–6.6] | 6.6  [5.8–7.5] | 5.8  [5.0–6.6] | 6.3  [5.5–7.2] | 5.4  [4.6–6.2] | 5.0  [4.1–6.0] | 6.0  [5.0–7.0] | 5.7  [4.7–6.7] | 6.0  [5.0–7.1] |
| ≥65 | 2.1  [1.6–2.6] | 3.0  [2.4–3.7] | 3.5  [2.8–4.1] | 3.6  [2.9–4.3] | 3.0  [2.5–3.5] | 4.7  [3.9–5.5] | 3.9  [3.2–4.5] | 4.7  [3.9–5.5] | 3.9  [3.1–4.7] | 4.2  [3.4–4.9] | 4.1  [3.5–4.8] | 3.5  [2.9–4.1] | 3.0  [2.4–3.7] | 4.3  [3.5–5.1] | 4.2  [3.3–5.1] | 4.0  [3.0–5.1] |
| Gender |  |  |  |  |  |  |  |  |  |  |  |  |  |  |  |  |
| Men | 5.4  [5.0–5.8] | 5.9  [5.5–6.2] | 5.9  [5.6–6.3] | 5.9  [5.5–6.3] | 6.1  [5.7–6.5] | 6.7  [6.3–7.1] | 6.1  [5.7–6.5] | 6.0  [5.6–6.5] | 5.9  [5.5–6.3] | 6.2  [5.8–6.7] | 6.1  [5.6–6.5] | 5.6  [5.2–6.0] | 6.0  [5.4–6.5] | 6.2  [5.7–6.8] | 6.1  [5.5–6.6] | 6.4  [5.8–7.1] |
| Women | 3.0  [2.8–3.3] | 3.1  [2.8–3.3] | 3.2  [2.9–3.4] | 3.4  [3.1–3.7] | 3.5  [3.2–3.8] | 4.1  [3.8–4.4] | 4.0  [3.7–4.4] | 4.8  [4.4–5.2] | 4.5  [4.2–4.9] | 4.5  [4.1–4.8] | 4.4  [4.0–4.7] | 4.5  [4.1–4.9] | 4.7  [4.2–5.1] | 4.7  [4.3–5.1] | 4.6  [4.2–5.0] | 4.9  [4.3–5.4] |
| Occupational social grade |  |  |  |  |  |  |  |  |  |  |  |  |  |  |  |  |
| ABC1 (more advantaged) | 2.7  [2.4–3.1] | 3.2  [2.9–3.5] | 3.2  [2.9–3.5] | 3.3  [2.9–3.6] | 3.7  [3.3–4.1] | 4.0  [3.7–4.4] | 3.7  [3.4–4.1] | 3.7  [3.4–4.1] | 3.4  [3.1–3.8] | 3.7  [3.4–4.1] | 3.9  [3.6–4.3] | 3.9  [3.6–4.3] | 4.2  [3.7–4.6] | 4.2  [3.8–4.6] | 4.1  [3.7–4.5] | 4.4  [3.9–4.8] |
| C2DE (less advantaged) | 5.1  [4.8–5.5] | 5.4  [5.1–5.7] | 5.7  [5.4–6.0] | 5.7  [5.4–6.1] | 5.7  [5.4–6.1] | 6.4  [6.1–6.8] | 5.9  [5.6–6.3] | 6.4  [6.0–6.8] | 6.4  [6.0–6.8] | 6.4  [6.0–6.8] | 6.1  [5.7–6.5] | 5.7  [5.3–6.1] | 6.2  [5.6–6.7] | 6.4  [5.9–6.9] | 6.3  [5.8–6.8] | 6.6  [6.0–7.3] |
| Region in England |  |  |  |  |  |  |  |  |  |  |  |  |  |  |  |  |
| North | 3.8  [3.4–4.2] | 4.0  [3.6–4.4] | 4.4  [4.0–4.8] | 4.3  [3.8–4.7] | 4.7  [4.3–5.2] | 5.5  [5.1–5.9] | 5.5  [5.1–6.0] | 6.0  [5.5–6.5] | 5.5  [5.0–6.0] | 5.7  [5.1–6.2] | 5.2  [4.7–5.7] | 5.3  [4.8–5.8] | 5.6  [4.9–6.3] | 5.6  [4.9–6.3] | 5.4  [4.8–6.0] | 5.6  [4.7–6.5] |
| Midlands | 4.7  [4.3–5.2] | 5.1  [4.6–5.5] | 5.0  [4.6–5.4] | 5.0  [4.6–5.5] | 5.2  [4.8–5.7] | 6.1  [5.6–6.7] | 4.5  [4.1–5.0] | 5.2  [4.6–5.8] | 5.0  [4.5–5.5] | 5.4  [4.9–5.9] | 5.8  [5.3–6.4] | 5.4  [4.9–5.9] | 5.8  [5.1–6.5] | 5.5  [5.0–6.1] | 5.6  [5.0–6.2] | 6.5  [5.7–7.4] |
| South | 4.1  [3.7–4.5] | 4.4  [4.1–4.8] | 4.6  [4.2–4.9] | 4.8  [4.5–5.2] | 4.9  [4.5–5.3] | 5.0  [4.6–5.4] | 5.2  [4.7–5.6] | 5.1  [4.6–5.5] | 5.2  [4.7–5.7] | 5.1  [4.6–5.6] | 4.9  [4.5–5.4] | 4.6  [4.2–5.0] | 5.0  [4.4–5.5] | 5.3  [4.8–5.8] | 5.2  [4.7–5.8] | 5.1  [4.6–5.7] |
| Current use of NRT |  |  |  |  |  |  |  |  |  |  |  |  |  |  |  |  |
| No | 4.3  [4.0–4.6] | 4.6  [4.4–4.9] | 4.8  [4.5–5.0] | 4.9  [4.6–5.1] | 4.9  [4.6–5.3] | 5.6  [5.3–5.9] | 5.2  [4.9–5.5] | 5.4  [5.1–5.7] | 5.3  [5.0–5.6] | 5.4  [5.1–5.8] | 5.3  [5.0–5.6] | 5.1  [4.8–5.3] | 5.4  [5.0–5.8] | 5.4  [5.0–5.7] | 5.3  [5.0–5.7] | 5.5  [5.1–6.0] |
| Yes | 3.7  [3.2–4.2] | 4.0  [3.5–4.5] | 3.9  [3.4–4.4] | 3.9  [3.4–4.5] | 4.3  [3.5–5.1] | 5.6  [4.8–6.5] | 4.5  [3.7–5.2] | 5.9  [4.9–6.9] | 4.9  [4.0–5.8] | 4.7  [3.8–5.6] | 5.4  [4.4–6.3] | 5.1  [4.1–6.1] | 5.3  [4.3–6.4] | 6.0  [4.8–7.3] | 5.9  [4.8–7.0] | 6.8  [5.0–8.7] |
| Current vaping^3^ |  |  |  |  |  |  |  |  |  |  |  |  |  |  |  |  |
| No | - | - | - | 4.8  [4.5–5.0] | 4.9  [4.6–5.3] | 5.7  [5.3–6.0] | 5.2  [4.9–5.5] | 5.5  [5.1–5.8] | 5.2  [4.9–5.5] | 5.3  [5.0–5.6] | 5.4  [5.0–5.7] | 5.2  [4.9–5.5] | 5.5  [5.1–5.9] | 5.6  [5.2–6.0] | 5.5  [5.1–5.9] | 5.8  [5.2–6.3] |
| Yes | - | - | - | 2.4  [1.1–3.6] | 3.6  [2.8–4.5] | 5.3  [4.6–6.1] | 4.9  [4.3–5.5] | 5.3  [4.7–6.0] | 5.4  [4.8–5.9] | 5.7  [5.0–6.4] | 4.9  [4.4–5.5] | 4.4  [3.8–5.0] | 5.1  [4.3–5.9] | 5.0  [4.3–5.6] | 5.1  [4.4–5.7] | 5.5  [4.8–6.3] |
| Non-daily | - | - | - | 1.5  [0.0–3.2] | 5.5  [3.5–7.6] | 5.2  [4.2–6.2] | 5.4  [4.3–6.4] | 5.9  [4.9–6.8] | 5.4  [4.4–6.4] | 5.7  [4.5–6.8] | 4.9  [3.9–5.9] | 4.6  [3.3–5.9] | 5.7  [4.2–7.2] | 5.7  [4.3–7.2] | 6.1  [4.2–8.1] | 7.0  [4.5–9.5] |
| Daily | - | - | - | 2.8  [1.1–4.5] | 2.7  [1.8–3.5] | 5.1  [4.0–6.2] | 4.6  [3.9–5.4] | 4.9  [4.1–5.7] | 5.1  [4.4–5.9] | 5.7  [4.8–6.6] | 5.0  [4.3–5.8] | 4.3  [3.6–5.0] | 5.0  [4.0–5.9] | 4.5  [3.7–5.4] | 5.3  [3.8–6.8] | 4.8  [3.7–6.0] |

^1^ Data are weighted means.

**^2^** Data for 2023 are based on January-September only.

**^3^** Vaping status has only been assessed since April 2011, so no data are available for 2008-2010 and data for 2011 are based on April-December only.

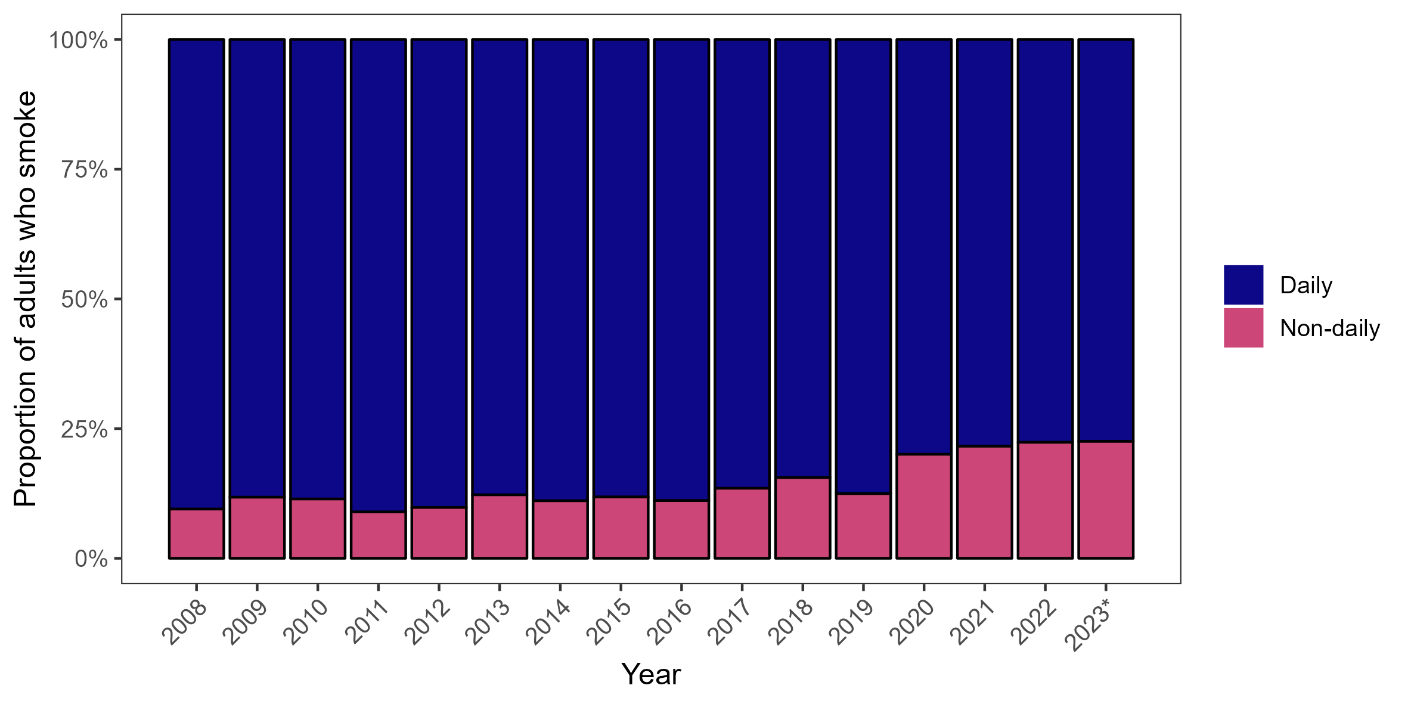

### **Figure S1. Frequency of smoking among adult** (≥18y) smokers in England, 2008 to 2023

The figure shows weighted data aggregated by year. Bars represent the proportion of smokers who smoke daily vs. non-daily.
* Data for 2023 are based on January to September only.

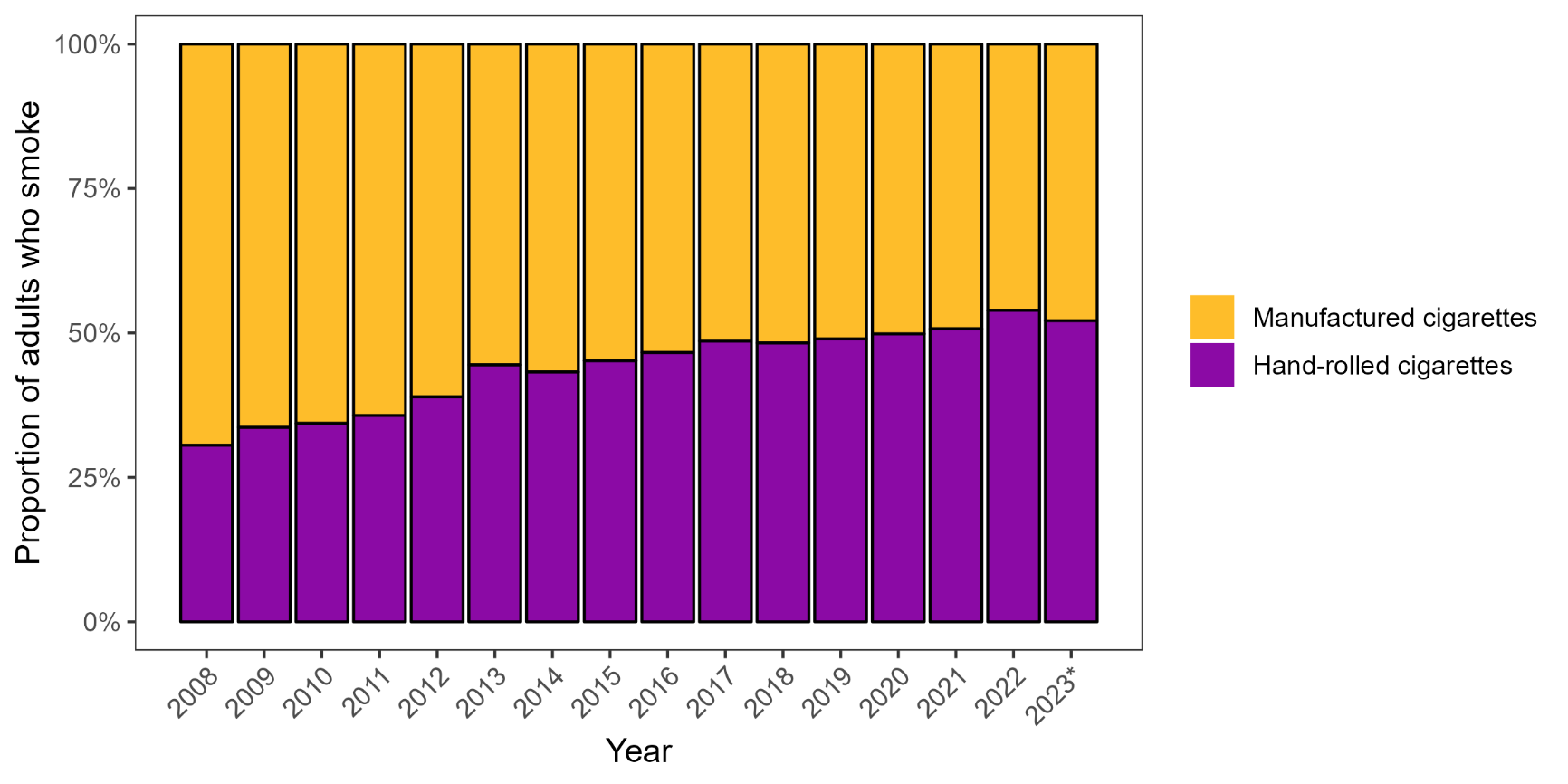

### **Figure S2. Main type of cigarette smoked among adult** (≥18y) smokers in England, 2008 to 2023

The figure shows weighted data aggregated by year. Bars represent the proportion of smokers who mainly smoke manufactured vs. hand-rolled cigarettes.
* Data for 2023 are based on January to September only.

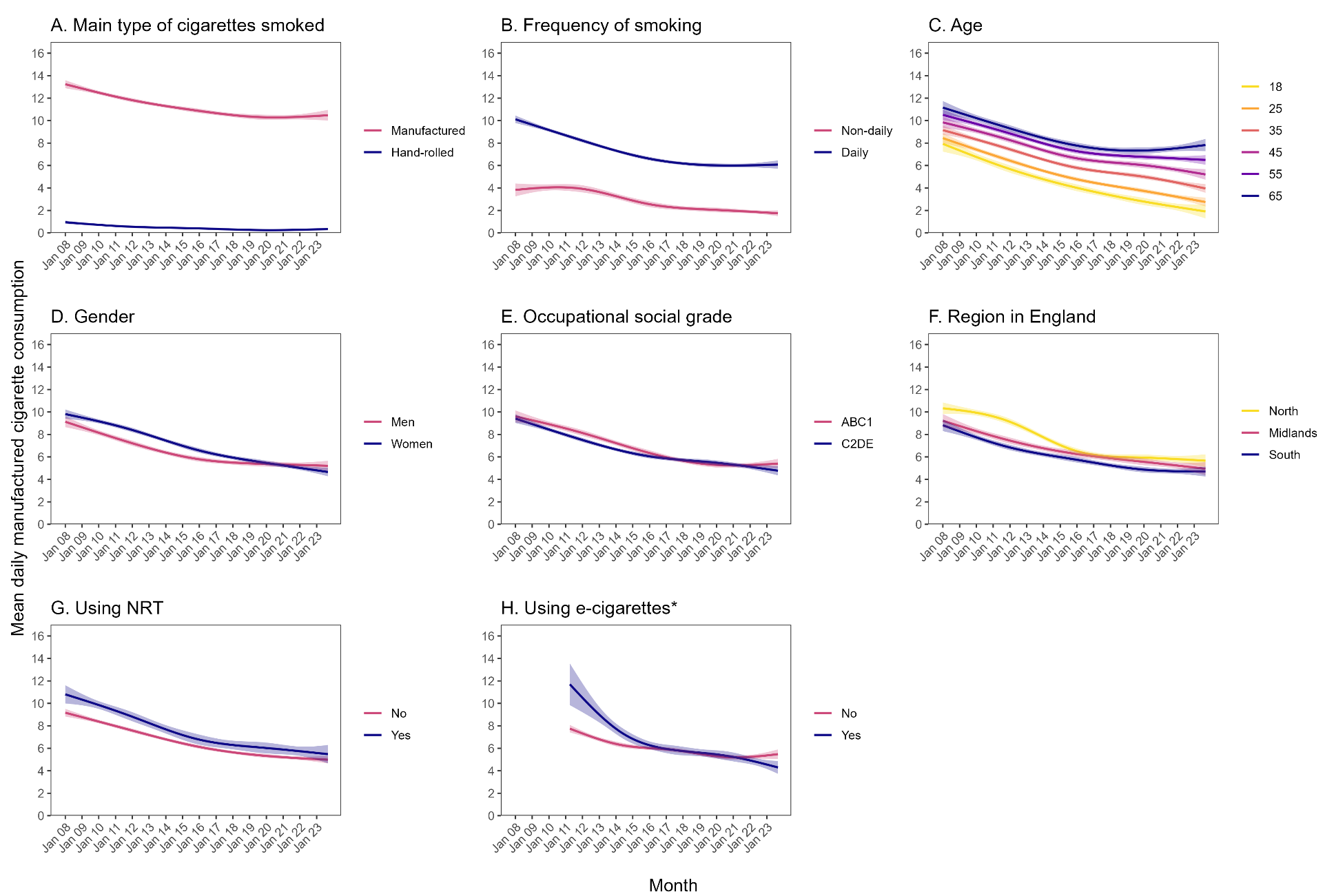

### **Figure S3. Time trends in daily manufactured cigarette consumption**

Panels show trends in daily manufactured cigarette consumption among adult (≥18y) smokers in England (January 2008 to September 2023) by (A) main type of cigarettes smoked, (B) frequency of smoking, (C) age, (D) gender, (E) occupational social grade, (F) region in England, (G) NRT use, and (H) vaping status. Lines represent modelled weighted mean daily cigarette consumption by monthly survey wave, modelled non-linearly using restricted cubic splines (five knots). Shaded bands represent 95% confidence intervals. * Vaping status has only been assessed since April 2011.

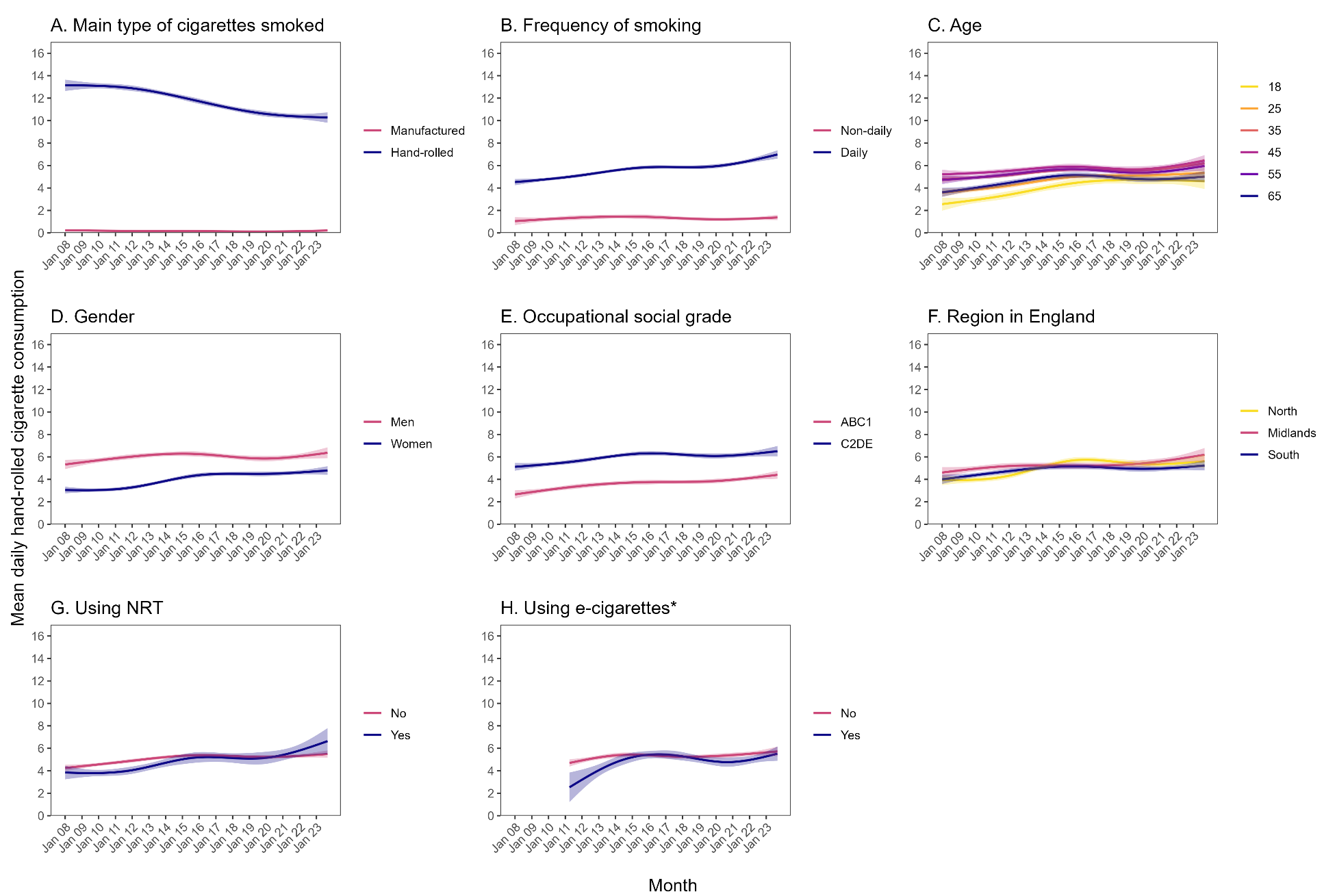

### **Figure S4. Time trends in daily hand-rolled cigarette consumption**

Panels show trends in daily hand-rolled cigarette consumption among adult (≥18y) smokers in England (January 2008 to September 2023) by (A) main type of cigarettes smoked, (B) frequency of smoking, (C) age, (D) gender, (E) occupational social grade, (F) region in England, (G) NRT use, and (H) vaping status. Lines represent modelled weighted mean daily cigarette consumption by monthly survey wave, modelled non-linearly using restricted cubic splines (five knots). Shaded bands **represent 95% confidence intervals**. * Vaping status has only been assessed since April 2011.

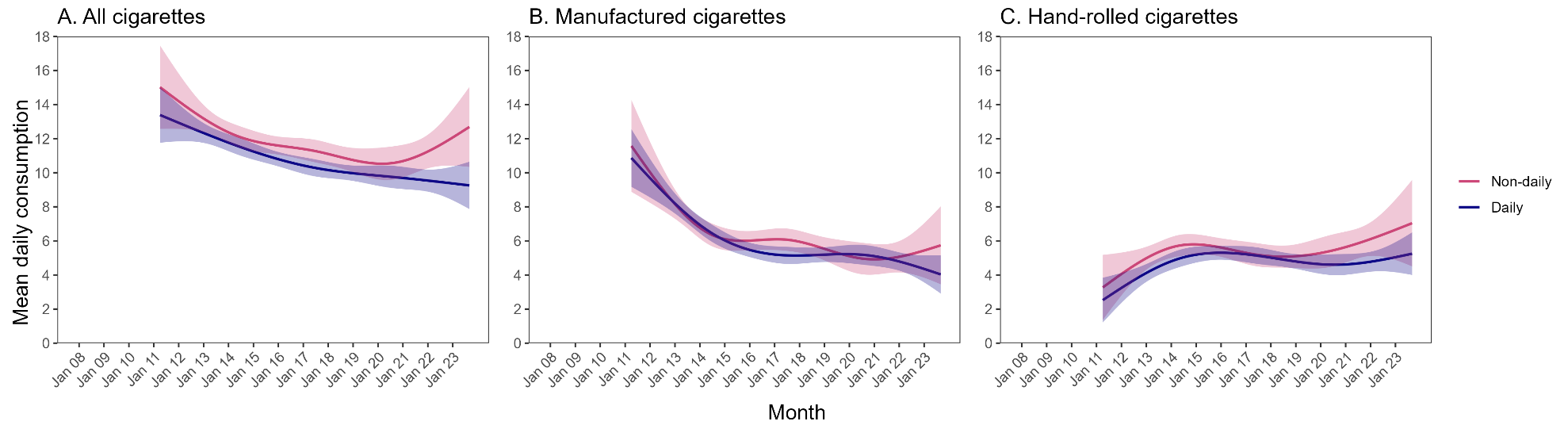

### **Figure S5. Time trends in daily cigarette consumption among current vapers**

Panels show trends in daily consumption of (A) all cigarettes, (B) manufactured cigarettes, and (C) hand-rolled cigarettes among adult (≥18y) smokers who currently vape in England (January 2008 to September 2023) by frequency of vaping (daily/non-daily). Lines represent modelled weighted mean daily cigarette consumption by monthly survey wave, modelled non-linearly using restricted cubic splines (five knots). Shaded bands represent 95% confidence intervals. Vaping status has only been assessed since April 2011.

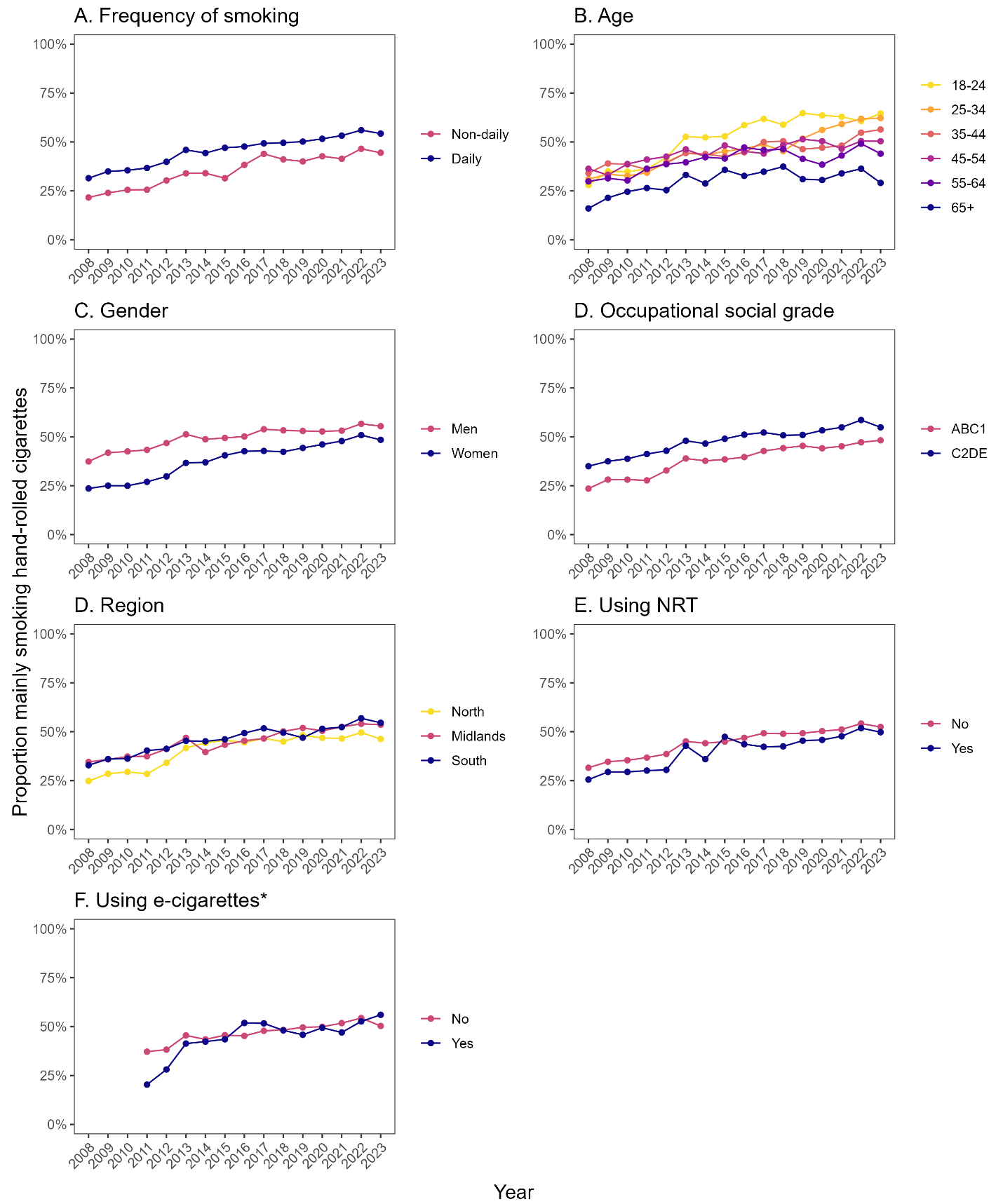

### **Figure S6. Time trends in the proportion mainly smoking hand-rolled cigarettes**

Panels show trends in the proportion of adult (≥18y) smokers who mainly smoked hand-rolled cigarettes, by (A) frequency of smoking, (B) age, (C) gender, (D) occupational social grade, (E) region in England, (F) NRT use, and (G) vaping status. Data shown are (unmodelled) weighted proportions, aggregated by survey year. * Vaping status has only been assessed since April 2011.
